# Association between Time-Weighted Remnant Cholesterol and Incident Cancer: A Population-Based Chinese Cohort Study

**DOI:** 10.1101/2024.03.07.24303903

**Authors:** Lifang Li, Nan Zhang, Yifan Yang, Hugo Hok Him Pui, Bosco Kwok Hei Leung, Oscar Hou In Chou, Carlin Chang, Abraham Ka Chung Wai, Gregory Lip, Gary Tse, Tong Liu, Jiandong Zhou

## Abstract

**Background:** Remnant cholesterol (RC) is becoming an increasingly well-recognized risk factor for cardiometabolic diseases. However, no study has explored the predictive role of RC in new-onset cancer. This study aimed to examine the associations between RC and time-weighted RC with incident cancer in the general population.

**Methods:** This was a retrospective population-based study enrolling patients attending family medicine clinics in Hong Kong between 1st January 2000 and 31st December 2003 with at least three RC measurements during follow-up visits. The primary outcome was new-onset cancer. The secondary outcome was cancer-related mortality. Multivariable Cox regression was used to evaluate associations between baseline RC and time-weighted RC with outcomes.

**Results:** A total of 75,342 adults (39.7% males, mean age: 62.5 years old) were included. During a median follow-up of 16.8 years, 8335 (11.1%) incident cancer and 4349 (5.7%) cancer-related deaths were observed. After adjusting for potential confounders, one mmol/L increased of time-weighted RC was associated with 41% and 62% higher risk of incident cancer (HR, 1.41; 95%CI, 1.26-1.57; p<0.0001) and cancer-related mortality (HR, 1.62; 95%CI, 1.43-1.85; p<0.0001), respectively. However, no significant association between baseline RC with risk of new-onset cancer (HR, 1.04; 95%CI, 0.82-1.31; p=0.768) and cancer-related mortality (HR, 0.85; 95%CI, 0.61-1.17; p=0.315) in the adjusted model. The association between time-weighted RC and incident cancer was significant regardless of age, gender, and remained consistent amongst those with baseline use of most cardiometabolic agents, as well as those complicated with most comorbidities.

**Conclusions:** Higher time-weighted RC was associated with increased risk of new-onset cancer and cancer-related mortality amongst the general population.

## Introduction

The growing global burden of cancer is rapidly exceeding the current cancer control capacity. Worldwide, an estimated 28.4 million new cancer cases are projected to occur in 2040, a 47% increase from the corresponding 19.3 million cases in 2020. Additionally, cancer has ranked as a leading cause of death, with expected 16 million deaths from cancer in 2040 globally.^1^ Therefore, it is of utmost importance to develop effective preventive strategies to facilitate global cancer control, which requires identification of potentially modifiable risk factors and determination of their contribution to cancer burden.^2^

Dyslipidemia has been considered as an important risk factor for cancer. However, previous studies regarding the association between dyslipidemia with cancer have mainly focused on other lipid profiles, such as LDL and triglyceride.^3, 4^ Remnant cholesterol (RC), also called triglyceride-rich lipoprotein (TRL) cholesterol, which comprises cholesterol carried in very-low-density lipoproteins (VLDL), chylomicron remnants, and intermediate-density lipoproteins, has been increasingly acknowledged as an important risk factor for several burdensome diseases, such as atherosclerotic cardiovascular disease (ASCVD),^5^ diabetes,^6^ and nonalcoholic fatty liver disease.^7^ However, as a highly burdensome disease, so far, no study has investigated the role of RC in the development of cancer. Therefore, this study aimed to examine the associations between RC level and incident cancer in the general population. Besides, to provide a dynamic picture of RC’s long-term effects, this study also focused on the associations between time-weighted RC with new-onset cancer.

## Methods

This retrospective study was approved by the Institutional Review Board of the University of Hong Kong/Hospital Authority Hong Kong West Cluster (Reference No. UW 20-250) and complied with the Declaration of Helsinki. Due to the retrospective design and use of deidentified data, the need for patient consent was waived.

### Data source

Data were acquired from the Clinical Data Analysis Reporting System (CDARS) of the Hong Kong Hospital Authority, a statutory body that manages all public hospitals and their affiliated outpatient and day care facilities in Hong Kong, covering approximately 90% of the population and being the most representative electronic medical database available in Hong Kong.^8^ CDARS prospectively collects patient information including, but not limited to, demographic data, selected laboratory tests, diagnoses, drug prescriptions, procedures, and episodes of hospital visits since 1993. CDARS encodes diagnoses using the International Classification of Diseases, ninth revision (ICD-9), as CDARS has not implemented ICD-10 codes to date.^9^ Mortality data were acquired from the linked Hong Kong Death Registry, a governmental registry of mortality data for Hong Kong citizens. CDARS and the linked Hong Kong Death Registry have been used extensively in research, with good coding accuracy and data completeness as demonstrated in previous studies.^10, 11^

### Study population

Patients aged 18 years old or above, without prior cancer history, who attended a family medicine clinic in Hong Kong between 1^st^ Jan 2000 and 31^st^ December 2003 were included. Those without baseline RC data and at least three valid RC tests during follow-up visits were excluded.

### Follow-up and Outcomes

All patients were followed up until 31st December 2019. The primary outcome of this study was new-onset cancer. The secondary outcome was cancer morality. ICD-9 codes for identifying site-specific cancer were listed in **Supplemental Table 1**.

### Data collection

The following data at baseline were collected: age, gender, Charlson’s comorbidity index, prior comorbidities, medication prescriptions, and selected laboratory results (kidney function, liver function, glucose and lipid profiles). All comorbidities were identified using ICD-9 codes, which were listed in **Supplemental Table 1**. Baseline and variability measures of blood pressure, glucose and lipid profiles were presented. Variability profiles were evaluated by SD and time-weighted measures. The following formula, 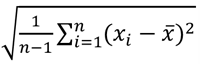, was used to calculated SD. Time-weighted measures was computed by the products of the sums of three consecutive measurements and the time interval, then divided by the total time interval.^12^ In addition, RC was calculated by total cholesterol minus LDL-C minus HDL-cholesterol (HDL-C).^13^ Besides, some calculated biomarkers, including the estimated glomerular filtration rate based on the abbreviated modification of diet in renal disease (AMDRD) formula,^14^ neutrophil-to-lymphocyte ratio (NLR),^15^ and triglyceride-glucose index (TGI) were also presented.^16^

### Statistical analysis

Descriptive statistics were used to summarize patients baseline clinical and biochemical characteristics. Continuous variables were presented as mean ± standard deviation (SD) or median with interquartile range (IQR) depending on their distribution. Categorical variables were presented as frequencies and percentages. Baseline RC and RC variability were assessed as both quartiles and continuous variable. Baseline characteristics of the included participants were compared according to the quantiles of RC and time-weighted RC, respectively. Kaplan-Meier curves were used to visualize the cumulative incidence of overall cancer and site-specific cancers across quartiles of baseline RC and time-weighted RC, respectively. Four separate Cox models were fitted hierarchically to estimate the adjusted hazard ratio (HR) and 95% CI for the associations between RC and time-weighted RC with outcomes. *A priori* subgroup analyses were performed using the fully adjusted Cox model with RC and time-weighted RC measures as continuous variables, according to age, gender, prior comorbidities, and medication prescriptions.

P value <0.05 was considered statistically significant. The statistical analysis was performed with RStudio software (Version 1.1.456) and Python (Version 3.6).

## Results

Altogether, 155,066 patients were identified. After applying the exclusion criteria, 75,342 adult patients with available baseline RC result and at least three RC results during follow-up visits were included in the final analysis (29,905 male [39.7%]; median age 62.5 years [interquartile range 51.5, 71.4 years]) (**Figure 1**). The baseline characteristics of included patients by RC and RC variability quartiles are summarized in **Table 1**.

**Figure 1.**
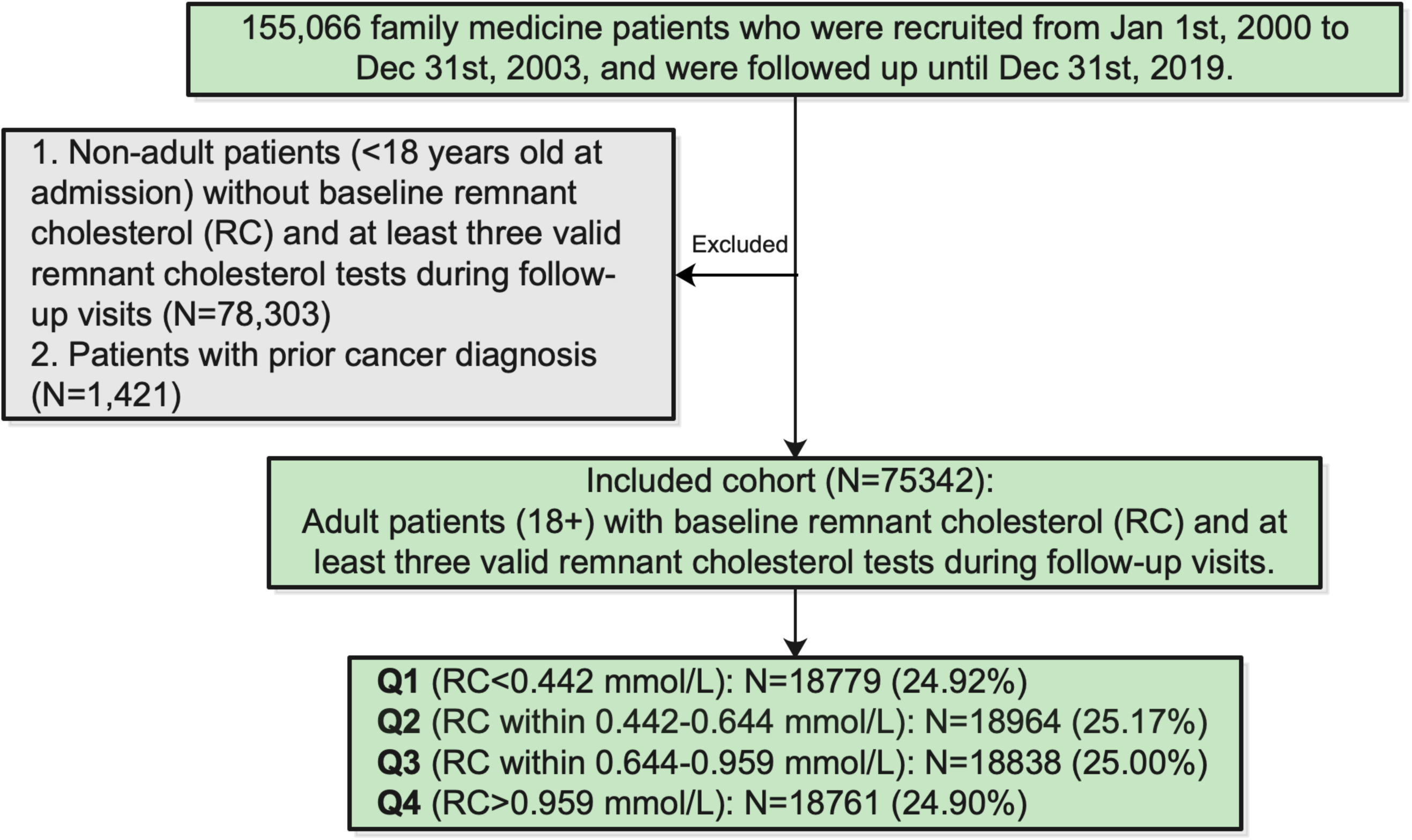
The flow diagram of the study.

**Table 1.** Baseline and clinical characteristics of family medicine patients stratified by quantiles of remnant cholesterol. SD: standard deviation; RC: remnant cholesterol; TIA: transient ischaemic attack; ACEI: angiotensin-converting enzyme inhibitors; ARB: angiotensin II receptor blockers. MDRD: modification of diet in renal disease.

| Characteristics | Baseline RC quantiles<br>Mean(SD);N or Count(%) |  |  |  |  |  | Time-weighted RC quantiles<br>Mean(SD);N or Count(%) |  |  |  |  |  |
| --- | --- | --- | --- | --- | --- | --- | --- | --- | --- | --- | --- | --- |
|  | Q1<br><0.44 mmol/L<br>(N=18779) | Q2<br>0.44-0.64<br>mmol/L<br>(N=18964) | Q3<br>0.64-0.96<br>mmol/L<br>(N=18838) | Q4<br>>0.96 mmol/L<br>(N=18761) | Total<br>(N=75342) | P<br>value | Q1<br><0.53 mmol/L<br>(N=18894) | Q2<br>0.53-0.70<br>mmol/L<br>(N=18820) | Q3<br>0.70-0.93<br>mmol/L<br>(N=18757) | Q4<br><0.93 mmol/L<br>(N=18871) | Total<br>(N=75342) | P<br>value |
| **Demographic** |  |  |  |  |  |  |  |  |  |  |  |  |
| Sex (Male) | 7697 (41.0) | 7519 (39.6) | 7245 (38.5) | 7444 (39.7) | 29905 (39.7) | <0.001 | 7896 (41.8) | 7363 (39.1) | 7120 (38.0) | 7526 (39.9) | 29905 (39.7) | <0.001 |
| Baseline age, years | 61.4<br>(49.8,71.2) | 62.9<br>(51.9,71.6) | 62.9<br>(52.2,71.3) | 62.6<br>(52.0,71.3) | 62.5<br>(51.5,71.4) | <0.001 | 62.9<br>(51.5,71.7) | 61.4<br>(51.2,70.1) | 61.3<br>(51.1,70.2) | 64.5<br>(52.2,73.5) | 62.5<br>(51.5,71.4) | <0.001 |
| Age larger than 65 | 7891 (42.0) | 8400 (44.3) | 8327 (44.2) | 8207 (43.7) | 32825 (43.6) | <0.001 | 8413 (44.5) | 7616 (40.5) | 7613 (40.6) | 9183 (48.7) | 32825 (43.6) | <0.001 |
| **Comorbidity** |  |  |  |  |  |  |  |  |  |  |  |  |
| Charlson's standard comorbidity index | 2.0 (0.0,3.0) | 2.0 (1.0,3.0) | 2.0 (1.0,3.0) | 2.0 (1.0,3.0) | 2.0 (1.0,3.0) | <0.001 | 2.0 (1.0,3.0) | 2.0 (1.0,3.0) | 2.0 (1.0,3.0) | 2.0 (1.0,3.0) | 2.0 (1.0,3.0) | <0.001 |
| Dyslipidemia | 188 (1.0) | 280 (1.5) | 348 (1.8) | 472 (2.5) | 1288 (1.7) | <0.001 | 259 (1.4) | 279 (1.5) | 368 (2.0) | 382 (2.0) | 1288 (1.7) | <0.001 |
| Diabetes mellitus | 1395 (7.4) | 1638 (8.6) | 2064 (11.0) | 2792 (14.9) | 7889 (10.5) | <0.001 | 1546 (8.2) | 1834 (9.7) | 2222 (11.8) | 2287 (12.1) | 7889 (10.5) | <0.001 |
| Hypertension | 8800 (46.9) | 9815 (51.8) | 10104 (53.6) | 10307 (54.9) | 39026 (51.8) | <0.001 | 9533 (50.5) | 9735 (51.7) | 9952 (53.1) | 9806 (52.0) | 39026 (51.8) | <0.001 |
| Heart failure | 557 (3.0) | 652 (3.4) | 784 (4.2) | 939 (5.0) | 2932 (3.9) | <0.001 | 591 (3.1) | 592 (3.1) | 735 (3.9) | 1014 (5.4) | 2932 (3.9) | <0.001 |
| Atrial fibrillation/Atrial flutter | 116 (0.6) | 120 (0.6) | 106 (0.6) | 104 (0.6) | 446 (0.6) | 0.688 | 122 (0.6) | 106 (0.6) | 102 (0.5) | 116 (0.6) | 446 (0.6) | 0.553 |
| Stroke/TIA | 92 (0.5) | 135 (0.7) | 139 (0.7) | 142 (0.8) | 508 (0.7) | 0.005 | 128 (0.7) | 113 (0.6) | 121 (0.6) | 146 (0.8) | 508 (0.7) | 0.207 |
| Ischemic heart disease | 322 (1.7) | 362 (1.9) | 395 (2.1) | 437 (2.3) | 1516 (2.0) | <0.001 | 345 (1.8) | 407 (2.2) | 391 (2.1) | 373 (2.0) | 1516 (2.0) | 0.107 |
| Liver diseases | 87 (0.5) | 68 (0.4) | 56 (0.3) | 58 (0.3) | 269 (0.4) | 0.028 | 75 (0.4) | 78 (0.4) | 65 (0.3) | 51 (0.3) | 269 (0.4) | 0.084 |
| Renal diseases | 26 (0.1) | 18 (0.1) | 31 (0.2) | 30 (0.2) | 105 (0.1) | 0.249 | 24 (0.1) | 17 (0.1) | 29 (0.2) | 35 (0.2) | 105 (0.1) | 0.084 |
| **Medication use** |  |  |  |  |  |  |  |  |  |  |  |  |
| ACEI/ARB | 1755 (9.3) | 2100 (11.1) | 2282 (12.1) | 2753 (14.7) | 8890 (11.8) | <0.001 | 1851 (9.8) | 2058 (10.9) | 2319 (12.4) | 2662 (14.1) | 8890 (11.8) | <0.001 |
| Antiplatelets | 1662 (8.9) | 1966 (10.4) | 2051 (10.9) | 2242 (12.0) | 7921 (10.5) | <0.001 | 1827 (9.7) | 1778 (9.4) | 1925 (10.3) | 2391 (12.7) | 7921 (10.5) | <0.001 |
| Lipid-lowering drugs | 1200 (6.4) | 1624 (8.6) | 1840 (9.8) | 2357 (12.6) | 7021 (9.3) | <0.001 | 1372 (7.3) | 1687 (9.0) | 1897 (10.1) | 2065 (10.9) | 7021 (9.3) | <0.001 |
| Statins and fibrates | 1353 (7.2) | 1825 (9.6) | 2207 (11.7) | 3075 (16.4) | 8460 (11.2) | <0.001 | 1506 (8.0) | 1893 (10.1) | 2348 (12.5) | 2713 (14.4) | 8460 (11.2) | <0.001 |
| Beta blockers | 2792 (14.9) | 3503 (18.5) | 4042 (21.5) | 4669 (24.9) | 15006 (19.9) | <0.001 | 3220 (17.0) | 3570 (19.0) | 4064 (21.7) | 4152 (22.0) | 15006 (19.9) | <0.001 |
| Non-steroidal anti-inflammatory drugs | 1604 (8.5) | 1890 (10.0) | 1964 (10.4) | 2136 (11.4) | 7594 (10.1) | <0.001 | 1759 (9.3) | 1706 (9.1) | 1830 (9.8) | 2299 (12.2) | 7594 (10.1) | <0.001 |
| Anti-diabetic drugs | 2860 (15.2) | 3483 (18.4) | 3996 (21.2) | 4782 (25.5) | 15121 (20.1) | <0.001 | 3104 (16.4) | 3329 (17.7) | 3922 (20.9) | 4766 (25.3) | 15121 (20.1) | <0.001 |
| **Calculated biomarkers** |  |  |  |  |  |  |  |  |  |  |  |  |
| eGFR (MDRD), mL/min/1.73m2 | 75.0<br>(64.4,86.4) | 72.9<br>(62.4,84.6) | 72.1<br>(61.5,83.9) | 71.1<br>(60.2,82.5) | 72.7<br>(61.9,84.3) | <0.001 | 73.4<br>(62.8,85.1) | 73.6<br>(63.1,84.9) | 73.1<br>(62.5,84.4) | 70.8<br>(59.3,82.6) | 72.7<br>(61.9,84.3) | <0.001 |
| Neutrophil-to-lymphocyte ratio | 2.3 (1.7,3.7) | 2.3 (1.7,3.6) | 2.2 (1.6,3.5) | 2.2 (1.6,3.5) | 2.2 (1.6,3.6) | 0.001 | 2.3 (1.6,3.6) | 2.2 (1.6,3.4) | 2.2 (1.6,3.4) | 2.3 (1.6,3.8) | 2.2 (1.6,3.6) | 0.004 |
| Triglyceride–glucose index | 6.7 (6.4,7.0) | 7.1 (6.9,7.3) | 7.4 (7.2,7.7) | 7.8 (7.5,8.2) | 7.3 (6.9,7.8) | <0.00<br>1 | 7.1 (6.7,7.5) | 7.2 (6.8,7.6) | 7.4 (7.0,7.8) | 7.5 (7.1,8.0) | 7.3 (6.9,7.8) | <0.00<br>1 |
| **kidney and liver function** |  |  |  |  |  |  |  |  |  |  |  |  |
| Creatinine, umol/L | 81.0<br>(70.0,96.0) | 83.0<br>(72.0,97.0) | 84.0<br>(72.0,98.0) | 85.0<br>(73.0,100.0) | 84.0<br>(72.0,98.0) | <0.00<br>1 | 83.0<br>(72.0,98.0) | 83.0<br>(72.0,96.0) | 83.0<br>(71.0,97.0) | 85.0<br>(73.0,100.0) | 84.0<br>(72.0,98.0) | <0.00<br>1 |
| Alkaline phosphatase, U/L | 74.0<br>(60.0,90.0) | 77.0<br>(63.0,94.0) | 78.0<br>(64.0,95.0) | 80.0<br>(66.0,97.0) | 78.0<br>(63.0,95.0) | <0.00<br>1 | 76.0<br>(62.0,93.0) | 77.0<br>(63.0,93.0) | 78.0<br>(64.0,95.0) | 79.0<br>(65.0,97.0) | 78.0<br>(63.0,95.0) | <0.00<br>1 |
| Aspartate transaminase, U/L | 19.0<br>(15.0,27.0) | 20.0<br>(15.0,28.0) | 21.0<br>(16.0,30.0) | 22.0<br>(17.0,31.0) | 21.0<br>(16.0,29.0) | <0.00<br>1 | 20.0<br>(15.0,27.0) | 21.0<br>(16.0,29.0) | 21.0<br>(16.0,30.0) | 21.0<br>(16.0,30.0) | 21.0<br>(16.0,29.0) | <0.00<br>1 |
| Alanine transaminase, U/L | 19.0<br>(14.0,28.0) | 20.0<br>(15.0,29.0) | 22.0<br>(15.0,31.0) | 23.0<br>(16.0,34.0) | 21.0<br>(15.0,31.0) | <0.00<br>1 | 21.0<br>(15.0,29.0) | 21.0<br>(15.0,31.0) | 22.0<br>(16.0,32.0) | 21.0<br>(15.0,31.0) | 21.0<br>(15.0,31.0) | <0.00<br>1 |
| Bilirubin, umol/L | 10.0 (7.0,13.0) | 9.6 (7.0,12.9) | 9.5 (7.0,13.0) | 9.2 (7.0,12.4) | 9.5 (7.0,13.0) | 0.002 | 9.9 (7.0,13.0) | 9.9 (7.0,13.0) | 9.3 (7.0,12.7) | 9.3 (7.0,12.6) | 9.5 (7.0,13.0) | 0.001 |
| **Glucose and blood pressure profiles** |  |  |  |  |  |  |  |  |  |  |  |  |
| Systolic blood pressure, mmHg | 136.0<br>(120.0,150.0) | 139.0<br>(125.0,153.0) | 140.0<br>(127.0,153.0) | 140.0<br>(127.0,153.0) | 139.0<br>(125.0,152.0) | <0.00<br>1 | 138.0<br>(123.0,152.0) | 139.0<br>(125.0,152.0) | 139.0<br>(126.0,152.0) | 139.0<br>(125.0,153.0) | 139.0<br>(125.0,152.0) | <0.00<br>1 |
| SD of systolic blood pressure | 12.9 (9.9,16.1) | 13.2<br>(10.4,16.6) | 13.3<br>(10.6,16.6) | 13.5<br>(10.7,16.8) | 13.2<br>(10.4,16.5) | <0.00<br>1 | 13.1<br>(10.3,16.4) | 13.2<br>(10.5,16.4) | 13.3<br>(10.5,16.4) | 13.3<br>(10.3,16.9) | 13.2<br>(10.4,16.5) | 0.001 |
| Diastolic blood pressure, mmHg | 75.0<br>(68.0,82.0) | 76.0<br>(69.0,84.0) | 77.0<br>(69.0,84.0) | 77.0<br>(70.0,85.0) | 76.0<br>(69.0,84.0) | <0.00<br>1 | 76.0<br>(68.0,83.0) | 76.0<br>(69.0,84.0) | 77.0<br>(69.0,84.0) | 76.0<br>(68.0,84.0) | 76.0<br>(69.0,84.0) | <0.00<br>1 |
| SD of diastolic blood pressure | 7.5 (6.0,9.2) | 7.8 (6.2,9.5) | 7.8 (6.3,9.5) | 7.9 (6.4,9.6) | 7.8 (6.2,9.5) | <0.00<br>1 | 7.7 (6.1,9.4) | 7.8 (6.3,9.4) | 7.8 (6.3,9.5) | 7.8 (6.1,9.6) | 7.8 (6.2,9.5) | 0.002 |
| HbA1C, % | 7.0 (6.2,8.0) | 7.1 (6.3,8.1) | 7.1 (6.4,8.2) | 7.4 (6.5,8.5) | 7.2 (6.3,8.3) | <0.00<br>1 | 7.1 (6.3,8.2) | 7.1 (6.3,8.3) | 7.2 (6.4,8.2) | 7.2 (6.4,8.4) | 7.2 (6.3,8.3) | 0.003 |
| SD of HbA1C | 0.5 (0.3,0.9) | 0.5 (0.3,0.9) | 0.5 (0.3,0.9) | 0.6 (0.3,1.0) | 0.5 (0.3,0.9) | <0.00<br>1 | 0.5 (0.3,0.9) | 0.5 (0.3,0.9) | 0.5 (0.3,0.9) | 0.6 (0.3,1.0) | 0.5 (0.3,0.9) | 0.001 |
| Time weighted mean HbA1C, % | 7.2 (6.7,7.8) | 7.2 (6.7,7.8) | 7.2 (6.7,7.8) | 7.2 (6.7,7.8) | 7.2 (6.7,7.8) | 0.077 | 7.2 (6.7,7.8) | 7.2 (6.7,7.8) | 7.3 (6.8,7.8) | 7.3 (6.7,7.8) | 7.2 (6.7,7.8) | 0.004 |
| Fasting glucose, mmol/L | 5.6 (5.0,7.2) | 5.8 (5.1,7.5) | 6.0 (5.2,7.8) | 6.2 (5.3,8.1) | 5.9 (5.2,7.7) | <0.00<br>1 | 5.8 (5.1,7.5) | 5.9 (5.1,7.7) | 6.0 (5.2,7.8) | 6.0 (5.2,7.7) | 5.9 (5.2,7.7) | <0.00<br>1 |
| SD of fasting glucose | 0.6 (0.3,1.6) | 0.7 (0.3,1.6) | 0.7 (0.3,1.7) | 0.8 (0.3,1.7) | 0.7 (0.3,1.6) | <0.00<br>1 | 0.7 (0.3,1.7) | 0.7 (0.3,1.6) | 0.7 (0.3,1.6) | 0.7 (0.3,1.7) | 0.7 (0.3,1.6) | 0.005 |
| Time weighted mean fasting glucose, mmol/L | 6.7 (5.6,7.9) | 6.8 (5.6,7.9) | 6.9 (5.7,8.1) | 7.0 (5.9,8.1) | 6.8 (5.7,8.0) | <0.00<br>1 | 6.8 (5.7,8.0) | 6.8 (5.6,7.9) | 6.9 (5.8,8.0) | 7.0 (6.0,8.2) | 6.8 (5.7,8.0) | <0.00<br>1 |
| **Lipid profiles and variability** |  |  |  |  |  |  |  |  |  |  |  |  |
| Triglyceride, mmol/L | 0.8 (0.7,0.9) | 1.2 (1.1,1.3) | 1.7 (1.5,1.9) | 2.5 (2.1,3.1) | 1.4 (1.0,1.9) | <0.00<br>1 | 1.1 (0.8,1.5) | 1.3 (0.9,1.7) | 1.5 (1.1,2.1) | 1.7 (1.2,2.5) | 1.4 (1.0,1.9) | <0.00<br>1 |
| SD of triglyceride | 0.2 (0.1,0.3) | 0.3 (0.2,0.4) | 0.4 (0.3,0.5) | 0.6 (0.4,0.9) | 0.3 (0.2,0.5) | <0.00<br>1 | 0.2 (0.2,0.4) | 0.3 (0.2,0.4) | 0.4 (0.3,0.6) | 0.5 (0.3,0.8) | 0.3 (0.2,0.5) | <0.00<br>1 |
| Time weighted mean triglyceride, mmol/L | 1.2 (1.0,1.4) | 1.3 (1.2,1.6) | 1.5 (1.3,1.7) | 1.7 (1.5,2.1) | 1.4 (1.2,1.7) | <0.00<br>1 | 1.2 (1.1,1.4) | 1.4 (1.2,1.5) | 1.6 (1.3,1.8) | 1.8 (1.4,2.2) | 1.4 (1.2,1.7) | <0.00<br>1 |
| Low-density lipoprotein, mmol/L | 2.9 (2.4,3.5) | 3.2 (2.6,3.8) | 3.3 (2.7,3.9) | 3.0 (2.4,3.7) | 3.1 (2.5,3.7) | <0.00<br>1 | 3.0 (2.5,3.6) | 3.1 (2.6,3.7) | 3.2 (2.6,3.8) | 3.1 (2.5,3.7) | 3.1 (2.5,3.7) | <0.00<br>1 |
| SD of low-density lipoprotein | 0.5 (0.3,0.7) | 0.5 (0.3,0.8) | 0.6 (0.4,0.8) | 0.6 (0.4,0.8) | 0.6 (0.3,0.8) | <0.00<br>1 | 0.5 (0.3,0.7) | 0.6 (0.4,0.8) | 0.6 (0.4,0.8) | 0.6 (0.3,0.8) | 0.6 (0.3,0.8) | <0.00<br>1 |
| Time weighted mean low-density lipoprotein, mmol/L | 2.7 (2.4,3.0) | 2.7 (2.4,3.0) | 2.7 (2.4,3.0) | 2.7 (2.4,3.0) | 2.7 (2.4,3.0) | <0.00<br>1 | 2.7 (2.5,3.1) | 2.7 (2.4,3.0) | 2.7 (2.4,3.0) | 2.6 (2.3,3.0) | 2.7 (2.4,3.0) | <0.00<br>1 |
| High-density lipoprotein, mmol/L | 1.5 (1.3,1.8) | 1.4 (1.1,1.6) | 1.2 (1.1,1.4) | 1.1 (1.0,1.3) | 1.3 (1.1,1.6) | <0.00<br>1 | 1.4 (1.1,1.7) | 1.3 (1.1,1.6) | 1.3 (1.1,1.5) | 1.2 (1.0,1.4) | 1.3 (1.1,1.6) | <0.00<br>1 |
| SD of high-density lipoprotein | 0.2 (0.1,0.2) | 0.1 (0.1,0.2) | 0.1 (0.1,0.2) | 0.1 (0.1,0.2) | 0.1 (0.1,0.2) | <0.00<br>1 | 0.2 (0.1,0.2) | 0.2 (0.1,0.2) | 0.1 (0.1,0.2) | 0.1 (0.1,0.2) | 0.1 (0.1,0.2) | <0.00<br>1 |
| Time weighted mean high-density lipoprotein, mmol/L | 1.4 (1.3,1.6) | 1.3 (1.2,1.5) | 1.3 (1.2,1.4) | 1.2 (1.1,1.4) | 1.3 (1.2,1.5) | <0.00<br>1 | 1.4 (1.3,1.6) | 1.3 (1.2,1.5) | 1.3 (1.2,1.4) | 1.2 (1.1,1.3) | 1.3 (1.2,1.5) | <0.00<br>1 |
| Total cholesterol, mmol/L | 4.8 (4.2,5.5) | 5.1 (4.5,5.8) | 5.3 (4.7,6.0) | 5.6 (5.0,6.4) | 5.2 (4.6,5.9) | <0.00<br>1 | 5.1 (4.5,5.8) | 5.2 (4.6,5.9) | 5.3 (4.7,6.0) | 5.4 (4.7,6.1) | 5.2 (4.6,5.9) | <0.00<br>1 |
| SD of total cholesterol | 0.5 (0.3,0.8) | 0.6 (0.4,0.8) | 0.7 (0.4,0.9) | 0.8 (0.5,1.0) | 0.6 (0.4,0.9) | <0.00<br>1 | 0.6 (0.4,0.8) | 0.6 (0.4,0.9) | 0.7 (0.4,0.9) | 0.6 (0.4,0.9) | 0.6 (0.4,0.9) | <0.00<br>1 |
| Time weighted mean total cholesterol, mmol/L | 4.7 (4.4,5.1) | 4.7 (4.4,5.1) | 4.8 (4.4,5.1) | 4.8 (4.5,5.2) | 4.7 (4.4,5.1) | <0.00<br>1 | 4.6 (4.2,4.9) | 4.7 (4.4,5.0) | 4.8 (4.5,5.1) | 5.0 (4.7,5.4) | 4.7 (4.4,5.1) | <0.00<br>1 |
| **Remnant cholesterol tests and variability** |  |  |  |  |  |  |  |  |  |  |  |  |
| Remnant cholesterol, mmol/L | 0.3 (0.3,0.4) | 0.5 (0.5,0.6) | 0.8 (0.7,0.9) | 1.3 (1.1,1.6) | 0.6 (0.4,1.0) | <0.00<br>1 | 0.5 (0.4,0.7) | 0.6 (0.4,0.8) | 0.7 (0.5,1.0) | 0.8 (0.5,1.3) | 0.6 (0.4,1.0) | <0.00<br>1 |
| Time weighted remnant cholesterol, mmol/L | 0.6 (0.4,0.8) | 0.6 (0.5,0.8) | 0.7 (0.6,0.9) | 0.9 (0.7,1.1) | 0.7 (0.5,0.9) | <0.00<br>1 | 0.4 (0.3,0.5) | 0.6 (0.6,0.7) | 0.8 (0.7,0.9) | 1.2 (1.0,1.8) | 0.7 (0.5,0.9) | <0.00<br>1 |

During a median follow-up of 16.8 (14.4, 17.9) years, 8335 (11.1%) patients developed incident cancer and 4349 (5.7%) died from cancer. Among the incident cancer cases, gastrointestinal cancer (N=3790, 5.0%) represented the most common type, followed by genitourinary cancer (N=2117, 2.8%), colorectal cancer (N=2051, 2.7%) and other types (**Supplemental Table 3 and 4**). The median baseline RC and time weighted RC of the overall cohort was 0.6 (0.4, 1.0) mmol/L and 0.7 (0.5, 0.9) mmol/L, respectively (**Table 1**). The relationship between RC with time-weighted RC, HDL-C, LDL-C, total cholesterol, and triglyceride on incident cancer were illustrated in **Supplementary** Figure 1.

### Association between baseline RC and outcomes

Cumulative incidence curves showed that groups with incrementally higher levels of remnant cholesterol had progressive higher incidences of overall cancer (**Figure 2A**). **Table 2** summarizes results of multivariable Cox regression models evaluating the associations between the RC level and the risk of incident cancer. After adjusting for demographics, comorbidities, medications, AMDRD, NLR, TGI, baseline and variability of blood pressure, glucose and other lipid profiles, no significant association was observed between baseline RC level with new-onset cancer (HR, 1.04; 95%CI, 0.82-1.31; p=0.768), or cancer-related mortality (HR, 0.85; 95%CI, 0.61-1.17; p=0.315). Results for the associations between RC with individual cancer types after adjusting for confounders were also in line with the primary analysis (**Table 2; Supplemental Figure 2**).

**Figure 2.**
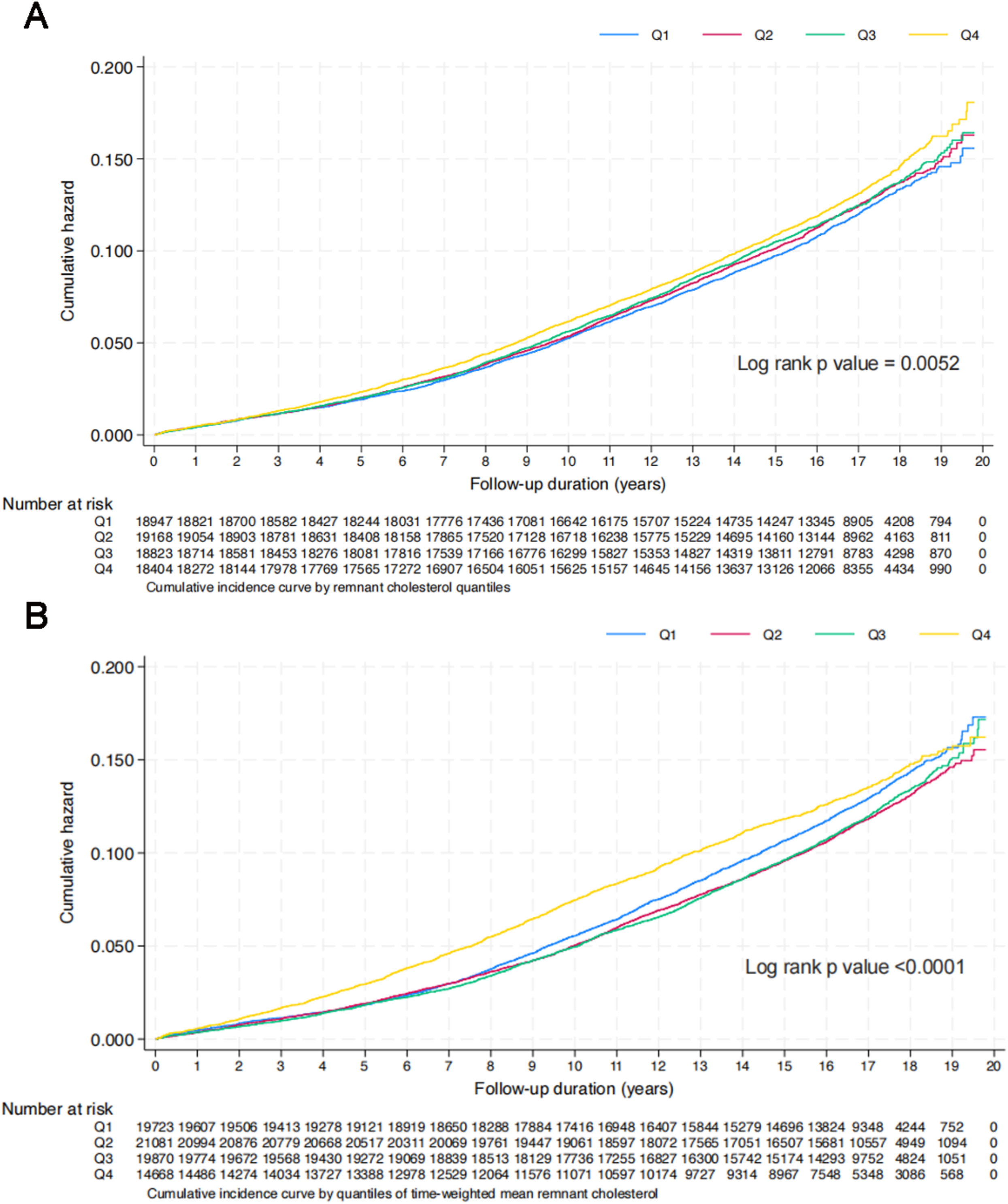
Cumulative incidence curves between (A) baseline remnant cholesterol and (B) time-weighted remnant cholesterol with incidence of overall cancer.

**Table 2.** Results of multivariate Cox analyses between baseline RC and time-weighted RC with outcomes. * for p≤ 0.05, ** for p ≤ 0.01, *** for p ≤ 0.001, Abbreviations: AMDRD, abbreviated modification of diet in renal disease; RC, remnant cholesterol; HR, hazard ratio.

| Models | Effects on RC<br>aHR [95% CI];P value | Effects of time-weighted RC<br>aHR [95% CI];P value |
| --- | --- | --- |
| **Model 1: Being adjusted for demographics** |  |  |
| Overall cancer | 1.07[1.03-1.11];0.0003*** | 1.02[0.99-1.05];0.1204 |
| Cancer mortality | 1.11[1.05-1.16];0.0001*** | 1.24[1.21-1.27];<0.0001*** |
| Lung cancer | 1.10[1.01-1.19];0.0214* | 1.13[1.07-1.19];<0.0001*** |
| Gastrointestinal cancer | 1.04[0.98-1.10];0.2246 | 1.03[0.99-1.07];0.1976 |
| Breast cancer | 1.18[1.08-1.28];0.0001*** | 0.95[0.87-1.03];0.2110 |
| Ovarian cancer | 1.15[0.86-1.55];0.3386 | 1.20[0.98-1.48];0.0778 |
| Prostate cancer | 0.94[0.83-1.08];0.3856 | 0.96[0.88-1.05];0.3372 |
| Genitourinary cancer | 1.03[0.95-1.11];0.4792 | 0.97[0.91-1.03];0.2569 |
| Colorectal cancer | 1.02[0.95-1.11];0.5595 | 0.98[0.93-1.04];0.5266 |
| Pancreas cancer | 1.19[1.03-1.38];0.0188* | 0.99[0.84-1.15];0.8517 |
| Bladder cancer | 1.10[0.95-1.28];0.1864 | 0.91[0.81-1.03];0.1370 |
| Liver cancer | 0.81[0.68-0.97];0.0211* | 1.10[1.00-1.21];0.0555 |
| **Model 2: Being adjusted for demographics, comorbidities, medications** |  |  |
| Overall Cancer | 1.05[1.01-1.09];0.0092** | 1.04[1.01-1.07];0.0045** |
| Cancer mortality | 1.07[1.01-1.13];0.0142* | 1.27[1.24-1.31];<0.0001*** |
| Lung cancer | 1.07[0.98-1.17];0.1078 | 1.15[1.09-1.21];<0.0001*** |
| Gastrointestinal cancer | 0.99[0.94-1.06];0.8638 | 1.05[1.01-1.09];0.0229* |
| Breast cancer | 1.14[1.06-1.23];0.0008*** | 0.95[0.87-1.04];0.2793 |
| Ovarian cancer | 1.12[0.86-1.46];0.4052 | 1.18[0.96-1.46];0.1094 |
| Prostate cancer | 0.86[0.74-0.99];0.0356* | 1.00[0.91-1.09];0.9494 |
| Genitourinary cancer | 1.00[0.92-1.09];0.9848 | 0.99[0.94-1.06];0.8574 |
| Colorectal cancer | 0.97[0.90-1.06];0.5423 | 1.00[0.94-1.06];0.9656 |
| Pancreas cancer | 1.20[1.02-1.42];0.0287* | 1.02[0.87-1.19];0.8202 |
| Bladder cancer | 1.05[0.89-1.24];0.5406 | 0.96[0.85-1.09];0.5538 |
| Liver cancer | 0.77[0.64-0.93];0.0061** | 1.13[1.02-1.25];0.0172* |
| **Model 3: Being adjusted for demographics, comorbidities, medications, AMDRD, calculated biomarkers, blood pressure** |  |  |
| Overall Cancer | 1.02[0.93-1.11];0.6660 | 1.26[1.20-1.32];<0.0001*** |
| Cancer mortality | 0.97[0.86-1.10];0.6764 | 1.49[1.42-1.57];<0.0001*** |
| Lung cancer | 1.06[0.88-1.27];0.5480 | 1.36[1.23-1.49];<0.0001*** |
| Gastrointestinal cancer | 0.93[0.81-1.07];0.3269 | 1.31[1.22-1.41];<0.0001*** |
| Breast cancer | 0.88[0.69-1.13];0.3214 | 1.06[0.88-1.26];0.5458 |
| Ovarian cancer | 1.61[1.04-2.50];0.0325* | 1.41[0.91-2.19];0.1267 |
| Prostate cancer | 1.03[0.77-1.38];0.8209 | 1.11[0.92-1.33];0.2782 |
| Genitourinary cancer | 1.03[0.86-1.23];0.7636 | 1.12[1.00-1.26];0.0566 |
| Colorectal cancer | 0.99[0.83-1.18];0.9290 | 1.25[1.13-1.38];<0.0001*** |
| Pancreas cancer | 1.26[0.91-1.75];0.1706 | 1.44[1.15-1.81];0.0016** |
| Bladder cancer | 1.03[0.73-1.43];0.8831 | 1.11[0.91-1.35];0.3218 |
| Liver cancer | 0.42[0.24-0.72];0.0016** | 1.33[1.11-1.59];0.0022** |
| **Model 4: Being adjusted for demographics, comorbidities, medications, AMDRD, calculated biomarkers, baseline and time-weighted blood pressure, glucose and lipid profiles** |  |  |
| Overall Cancer | 1.04[0.82-1.31];0.7677 | 1.41[1.26-1.57];<0.0001*** |
| Cancer mortality | 0.85[0.61-1.17];0.3153 | 1.62[1.43-1.85];<0.0001*** |
| Lung cancer | 0.71[0.40-1.27];0.2451 | 1.60[1.28-2.00];<0.0001*** |
| Gastrointestinal cancer | 0.97[0.70-1.35];0.8754 | 1.39[1.18-1.64];0.0001*** |
| Breast cancer | 1.50[0.89-2.52];0.1259 | 1.12[0.74-1.70];0.5904 |
| Ovarian cancer | 1.46[0.35-6.04];0.5990 | 2.71[1.23-5.95];0.0133* |
| Prostate cancer | 0.98[0.49-1.93];0.9440 | 1.27[0.91-1.77];0.1682 |
| Genitourinary cancer | 0.91[0.54-1.54];0.7375 | 1.27[1.00-1.61];0.0483* |
| Colorectal cancer | 1.23[0.84-1.82];0.2912 | 1.47[1.21-1.78];0.0001*** |
| Pancreas cancer | 0.47[0.13-1.73];0.2564 | 0.45[0.09-2.39];0.3512 |
| Bladder cancer | 1.04[0.38-2.84];0.9388 | 0.73[0.31-1.73];0.4705 |
| Liver cancer | 0.79[0.30-2.07];0.6259 | 0.86[0.38-1.95];0.7213 |

### Association between time-weighted RC and outcomes

Cumulative incidence curves showed a significant association between higher RC variability with increased risk of incident cancer (**Figure 2B**). In the fully adjusted model, one mmol/L increased of time-weighted RC was associated with 41% and 62% higher risk of incident cancer (HR, 1.41; 95%CI, 1.26-1.57; p<0.0001) and cancer-related mortality (HR, 1.62; 95%CI, 1.43-1.85; p<0.0001), respectively. For individual cancer types, higher time-weighted RC was associated with significantly increased risk of lung cancer (HR, 1.60, 95%CI, 1.28-2.00; p<0.0001), gastrointestinal cancer (HR, 1.39; 95%CI, 1.18-1.64; p=0.0001), colorectal cancer (HR, 1.47; 95%CI, 1.21-1.78; P=0.0001), ovarian cancer (HR, 2.707; 95%CI, 1.231-5.954; P=0.0133), and genitourinary cancer (HR, 1.27; 95%CI, 1.00-1.61; p=0.0483), but not for breast cancer (HR, 1.12; 95%CI 0.74-1.70; p=0.5904), prostate cancer (HR, 1.27; 95%CI, 0.91-1.77; p=0.1682), pancreatic cancer (HR, 0.45; 95%CI, 0.09-2.39; p=0.3512), liver cancer (HR, 0.86; 95%CI, 0.38-1.95; p=0.7213), or bladder cancer (HR, 0.73; 95%CI, 0.31-1.73; p=0.4705) **(Table 2; Supplemental Figure 3).**

### Subgroup analyses

Subgroup analyses identified a significant association between time-weighted RC and incident cancer both in females (HR 1.07; 95% CI 1.03-1.11; P=0.001) and males (HR 1.10; 95% CI 1.06-1.15; P<0.001) (**Figure 3**). In subgroup analysis by age, the association between time-weighted RC and cancer was significant among both the younger (<65 years, HR 1.07; 95% CI 1.01-1.12; P=0.013) and older (>65 years, HR 1.05; 95% CI 1.02-1.09; P = 0.004) individuals (**Figure 3**).

**Figure 3.**
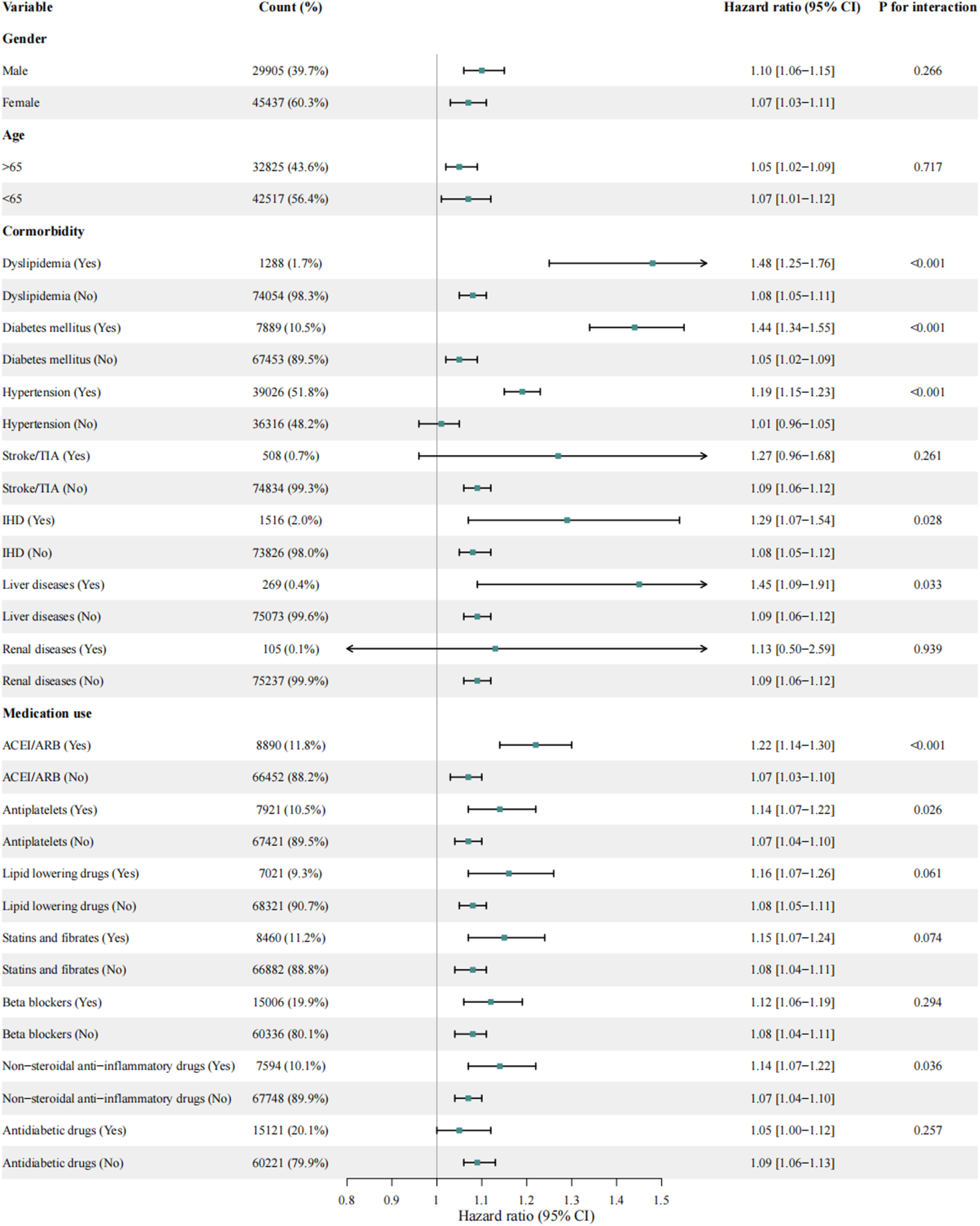
Forest plot of subgroup analyses between time-weighted RC with cancer.

Further analyses were performed according to prior comorbidities, which suggested a significant association between time-weighted RC and cancer regardless of the presence of baseline dyslipidemia, diabetes mellitus, ischemic heart disease, and liver disease (**Figure 3**). However, the association between time-weighted RC with cancer was more prominent among those complicated with baseline hypertension (HR, 1.19; 95%CI 1.15-1.23; P<0.001), but not for those without (HR, 1.01; 95%CI 0.96-1.05; P=0.812), with a P for interaction < 0.001. In subgroup analyses according to medication use, higher time-weighted RC was consistently associated with increased risk of cancer, regardless of the baseline usage of angiotensin-converting enzyme inhibitor (ACEI)/angiotensin receptor blocker (ARB), beta-blockers, antiplatelets, statins and fibrates, overall lipid-lowering drugs, and non-steroidal anti-inflammatory agents. Compared to patients treated with anti-diabetic drugs, those without exposure to anti-diabetic drugs presented with a significant association between time-weighted RC with cancer (HR, 1.09; 95%CI 1.06-1.13; P<0.001), whereas the interaction did not reach the traditional significance (P_for interaction_=0.257) (**Figure 3**).

Results from subgroup analyses between RC and cancer were generally consistent with those observed between time-weighted RC with cancer (**Supplemental Figure 4**). Notably, higher RC level was associated with significantly increased risk of new-onset cancer in female individuals (HR 1.17; 95% CI 1.11-1.22; P<0.001), but not for male (HR 0.96; 95% CI 0.90-1.02; P=0.199), with a p for interaction < 0.001. Besides, Higher RC level was associated with significantly increased risk of cancer among those without medication history of statins and fibrates (HR, 1.10; 95%CI 1.06-1.15, p<0.001), but not for those exposure to statins and fibrates (HR, 0.96; 95%CI 0.88-1.05, p=0.363), with a p for interaction of 0.004.

## Discussion

In this population-based cohort study with over 16 years of follow-up, several key findings were noted. First, higher time-weighted RC, but not single time point RC, was significantly associated with increased risks of new-onset cancer, especially for lung cancer, gastrointestinal cancer, genitourinary cancer, and ovarian cancer. Second, the association between time-weighted RC and incident cancer was significant regardless of age and gender. The association remained consistent amongst those with baseline use of most cardiometabolic agents, as well as those complicated with most comorbidities. Last, higher time-weighted RC was associated with significantly increased risk of cancer-related mortality, but not for single RC measurement. To our knowledge, the present study is the first to demonstrate the effects of RC and time-weighted RC in the development of cancer and cancer-related mortality.

### Comparison with previous studies

Previous preclinical studies have suggested a role of cholesterol in cancer, with several demonstrating that cholesterol homeostasis genes can modulate cancer development.^4^ In addition, some epidemiologic studies also identified an association between higher serum cholesterol and LDL level with increased risk of incident cancer.^20, 21^ In line with the prior studies, our study observed that higher time-weighted RC was significantly associated with increased risk of new-onset cancer in the general population, which adds to the existent evidence that time-weighted RC may be a potential tool for cancer risk assessment. In the present study, the predictive role of time-weighted RC was only observed in lung cancer, colorectal cancer, ovarian cancer, overall gastrointestinal cancer and genitourinary cancer. The discrepancy of the predictive role of lipid profile across various site-specific cancers has also been observed in other studies.^21^ Prior investigation has demonstrated that intracellular cholesterol homeostasis varies among different cancer types,^22^ therefore, whether RC plays differing roles in various cancer type needs further exploration. In addition, future large-scale study to reveal the genetic architecture of RC and nonlinear Mendelian randomization study to explore the causal association between RC and cancel are needed.

Though no study has addressed the association between RC and incident cancer, several observational studies have explored the relationship between RC and cancer-related mortality but yielded controversial results. Both Wadström *et al.*^23^ and Bonfiglio *et al.*^24^ have focused on the association between baseline RC levels with cancer mortality, whereas they failed to observe a significant correlation. In agreement with Wadström *et al.*^23^ and Bonfiglio *et al.*^24^, no significant association between baseline RC and cancer-related mortality was identified in the present study, whereas we found a significant association between time-weighted RC with cancer-related mortality, regardless of age, gender, and presence of most comorbidities and medication history. Time-weighted measurement, an index of homeostasis that takes into account the time spent at every single test, has been demonstrated to be superior to and more robust than static indices.^25, 26^ Therefore, by using the time-weighted measurement, our study could provide a dynamic picture of RC’s homeostasis on cancer risk and cancer mortality. Interestingly, in the study by Tian *et al.*, during a median follow-up of 3.6 years, an association between increased baseline RC and reduced risk of overall, liver and stomach cancer-related morality was observed.^27^ However, the prior study may be limited by the relatively short follow-up and the possibility of reverse causation.^27^ For example, liver cancer is usually accompanied by cirrhosis before its onset, which lowers cholesterol levels before the onset of liver cancer.^27, 28^

### Potential Underlying Mechanisms

The mechanisms underlying the associations between time-weighted RC with incident cancer and cancer mortality have not been addressed before. Based on current studies regarding other lipid traits with cancer, several hypotheses may help explain the observed associations. First of all, altered lipid metabolism is known to be a prominent metabolic alterations in cancer. Enhanced synthesis or uptake of lipids contributes to rapid cancer cell growth and tumor formation. Altered cholesterol metabolism has been demonstrated to contribute to various aspects of carcinogenesis, such as structural functions as components of cellular membranes,^29^ controlling the communication between cancer and immune cells within the cancer microenvironment,^30^ activity of multiple signaling pathways that are directly carcinogenic.^20, 31^ In addition, RC increases the production of reactive oxygen species and induces inflammatory response, both of which has been considered to play a critical role in the cancer development and progression.^32, 33^ Furthermore, lipid plays a major role in regulation of the processes that initiate cell dissemination and metastasis formation.^29, 34^ Metastasis is the prime cause of cancer-related deaths, which could help explain the association between time-weighted RC with cancer mortality observed in the present study. Future studies are clearly needed to investigate the mechanisms underlying the effects of RC on cancer development.

### Clinical Implications and the future

RC has been increasingly considered as a substantial risk factor for cardiometabolic diseases.^35^ Our findings add to the evidence that RC may also be a novel modifiable risk factor for new-onset cancer and cancer-related mortality. One of strengthens of using family medicine cohort is that population in our study generally represents the least unwell patients that one would encounter in daily clinical practice. Given the readily available and low cost of lipid testing, time-weighted RC has the potential to achieve widespread clinical use with minimal interference of general medical practices.^20^ Our findings suggest that including time-weighted RC in general medical assessments could allow not only stratification of cardiometabolic risk, but also cancer risk. Future researches are warranted to evaluate the effects of RC homeostasis management, such as behavioral or pharmacological intervention, on the prevention of cancer risk.

### Limitations

First, given the retrospective nature of this study, there might be unmeasured and residual confounders which have not been accounted for. Nonetheless, we have adjusted for a range of well-established risk factors in the multivariable Cox regression analyses, which should account for most potential confounding factors pertinent to our outcomes. Second, this study only included participants from Hong Kong; therefore, the findings may not be generalizable to other populations. Future studies evaluating the association between RC and incident cancer among individuals of other races are needed. In addition, causation relationship between RC and cancer using Mendelian randomization in Eastern Asian should be further investigated to testify our result. Last, there is inherent information bias due to under-coding, coding errors, and missing data. Nevertheless, previous studies have demonstrated good coding accuracy and data completeness in CDARS.^9, 10^

## Conclusions

Higher time-weighted RC was associated with increased risk of new-onset cancer and cancer-related mortality amongst the general population. Time-weighted RC may be considered as a potential tool for cancer risk assessment and optimization of RC homeostasis may potentially help prevent cancer development.

## Data Availability

Data are not available, as the data custodians (the Hospital Authority and the Department of Health of Hong Kong SAR) have not given permission for sharing due to patient confidentiality and privacy concerns. Local academic institutions, government departments, or nongovernmental organizations may apply for the access to data through the Hospital Authority's data sharing portal (https://www3.ha.org.hk/data).

## Ethical approval statement

This study was approved by the Institutional Review Board of the University of Hong Kong/Hospital Authority Hong Kong West Cluster (HKU/HA HKWC IRB) (UW-20-250) and complied with the Declaration of Helsinki.

## Guarantor Statement

All authors approved the final version of the manuscript. GT is the guarantor of this work and, as such,

had full access to all the data in the study and takes responsibility for the integrity of the data and the accuracy of the data analysis.

## Author contributions

Data analysis: LL, NZ, JZ

Data review: LL, NZ, GT, JZ

Data acquisition: HHHP, BKHL, AKCW

Data interpretation: LL, NZ, CC, JZ, GT

Critical revision of manuscription: AKCW, CC, TL, GL, GT, JZ

Supervision: TL, GT, JZ

Manuscript writing: LL, NZ, GT, JZ

Manuscript revision: LL, NZ, AKCW, TL, GL, GT, JZ

## Acknowledgements

All the authors and colleagues from the Hospital Authority for providing de-identified clinical data are equally thanked for their contributions to this research. Special thanks to the support of the National Natural Science Foundation of China (82170327, 82370332 to TL) and the Tianjin Key Medical Discipline (Specialty) Construction Project (TJYXZDXK-029A).

