## Supplementary Appendix for "Association between Time-Weighted Remnant Cholesterol and Incident Cancer: A Population-Based Chinese Cohort Study"

**Supplementary Table 1. The International Classification of Diseases (ICD)-9 codes for diagnoses and ICD-10 codes for outcomes.**

| |
|---|
| Diabetes mellitus 250 250.01 250.02 250.03 250.1 250.11 250.12 250.13 250.2 250.21 250.22 250.23 250.3 250.31 250.32 250.33 250.4 250.41 250.42 250.43 250.5 250.51 250.52 250.53 250.6 250.61 250.62 250.63 250.7 250.71 250.72 250.73 250.8 250.81 250.82 250.83 250.9 250.91 250.92 250.93 |
| Hypertension 401 401.1 401.9 402 402.01 402.1 402.11 402.9 402.91 403 403.01 403.1 403.11 403.9 403.91 404 404.01 404.02 404.03 404.1 404.11 404.12 404.13 404.9 404.91 404.92 404.93 405 405.01 405.09 405.1 405.11 405.19 405.9 405.91 405.99 437.2 |
| Heart failure 428 428 428.1 428.2 428.2 428.21 428.22 428.23 428.3 428.3 428.31 428.32 428.33 428.4 428.4 428.41 428.42 428.43 428.9 398.91 402.01 402.11 402.91 404.01 404.03 404.11 404.13 404.91 404.93 |
| Chronic kidney disease 585.1 585.2 585.3 585.4 585.5 585.6 585.9 |
| Stroke/transient ischemic attack 435 435.1 435.2 435.3 435.8 435.9 433.81 433.91 434 436 437 437.1 433.31 433.01 434.01 434.1 434.11 434.9 434.91 437.2 437.3 437.4 437.5 437.6 437.7 437.8 437.9 |
| Ischemic heart disease 410.01 410.02 410.1 410.11 410.12 410.2 410.21 410.22 410.3 410.31 410.32 410.4 410.41 410.42 410.5 410.51 410.52 410.6 410.61 410.62 410.7 410.71 410.72 410.8 410.81 410.82 410.9 410.91 410.92 411 411.1 411.8 411.81 411.89 413 413.1 413.9 414 414.01 414.02 414.03 414.04 414.05 414.06 414.07 414.1 414.11 414.12 414.19 414.2 414.3 414.4 414.8 414.9 410 412 |
| Cancer 140 141 142 143 144 145 146 147 148 149 150 151 152 153 154 155 156 157 158 159 160 161 162 163 164 165 170 171 172 173 174 175 176 179 180 181 182 183 184 185 186 187 188 189 190 191 192 193 194 195 196 197 198 199 200 201 202 203 204 205 206 207 208 209 |
| Dyslipidemia 272 272.0 272.1 272.2 272.3 272.4 |
| Cancer mortality (ICD-10 codes) I00-I09, I11, I13, I20-I51 |

**Supplementary Table 2. The outcome of patients in the study cohorts of four quartiles remnant cholesterol.**

Q: quantile; SD: standard deviation

| Characteristics | Q1 (N=18779) Mean(SD);N or Count(%) | Q2 (N=18964) Mean(SD);N or Count(%) | Q3 (N=18838) Mean(SD);N or Count(%) | Q4 (N=18761) Mean(SD);N or Count(%) | Total (N=75342) Mean(SD);N or Count(%) | P value |
|---|---|---|---|---|---|---|
| Overall cancer | 1995 (10.6) | 2090 (11.0) | 2082 (11.1) | 2168 (11.6) | 8335 (11.1) | 0.039 |
| Time to cancer, year | 16.8 (15.1, 17.9) | 16.8 (14.6, 17.9) | 16.8 (14.4, 17.9) | 16.7 (13.7, 17.9) | 16.8 (14.4, 17.9) | 0.004 |
| Lung cancer | 391 (2.1) | 398 (2.1) | 429 (2.3) | 422 (2.2) | 1640 (2.2) | 0.442 |
| Time to lung cancer, year | 17.0 (15.8, 17.9) | 17.0 (15.4, 17.9) | 17.0 (15.3, 18.0) | 16.9 (15.0, 18.0) | 17.0 (15.3, 18.0) | 0.054 |
| Gastrointestinal cancer | 951 (5.1) | 972 (5.1) | 927 (4.9) | 940 (5.0) | 3790 (5.0) | 0.827 |
| Time to gastrointestinal cancer, year | 16.9 (15.4, 17.9) | 16.9 (15.2, 17.9) | 16.9 (15.2, 17.9) | 16.8 (14.5, 18.0) | 16.9 (15.2, 17.9) | 0.054 |
| Breast cancer | 298 (1.6) | 317 (1.7) | 366 (1.9) | 389 (2.1) | 1370 (1.8) | 0.001 |
| Time to Breast cancer, year | 17.0 (15.6, 17.9) | 16.9 (15.3, 17.9) | 16.9 (15.2, 17.9) | 16.8 (14.6, 18.0) | 16.9 (15.2, 17.9) | 0.009 |
| Ovarian cancer | 22 (0.1) | 38 (0.2) | 32 (0.2) | 31 (0.2) | 123 (0.2) | 0.25 |
| Time to ovarian cancer, year | 17.0 (15.9, 17.9) | 17.0 (15.4, 17.9) | 17.0 (15.3, 18.0) | 16.9 (15.1, 18.0) | 17.0 (15.3, 18.0) | 0.056 |
| Prostate cancer | 230 (1.2) | 227 (1.2) | 215 (1.1) | 190 (1.0) | 862 (1.1) | 0.22 |
| Time to prostate cancer, year | 17.0 (15.7, 17.9) | 17.0 (15.3, 17.9) | 17.0 (15.3, 18.0) | 16.9 (14.9, 18.0) | 16.9 (15.3, 18.0) | 0.081 |
| Genitourinary cancer | 501 (2.7) | 559 (2.9) | 538 (2.9) | 519 (2.8) | 2117 (2.8) | 0.394 |
| Time to genitourinary cancer, year | 16.9 (15.5, 17.9) | 16.9 (15.3, 17.9) | 16.9 (15.2, 17.9) | 16.9 (14.6, 18.0) | 16.9 (15.2, 17.9) | 0.033 |
| Colorectal cancer | 494 (2.6) | 529 (2.8) | 516 (2.7) | 512 (2.7) | 2051 (2.7) | 0.815 |
| Time to colorectal cancer, year | 17.0 (15.6, 17.9) | 16.9 (15.3, 17.9) | 16.9 (15.2, 17.9) | 16.9 (14.7, 18.0) | 16.9 (15.2, 17.9) | 0.032 |
| Pancreas cancer | 68 (0.4) | 70 (0.4) | 78 (0.4) | 76 (0.4) | 292 (0.4) | 0.808 |
| Time to pancreas cancer, year | 17.0 (15.9, 17.9) | 17.0 (15.4, 17.9) | 17.0 (15.3, 18.0) | 16.9 (15.1, 18.0) | 17.0 (15.3, 18.0) | 0.069 |
| Time to metastatic solid tumour, year | 17.0 (15.9, 17.9) | 17.0 (15.4, 17.9) | 17.0 (15.3, 18.0) | 16.9 (15.0, 18.0) | 17.0 (15.3, 18.0) | 0.053 |
| Bladder cancer | 114 (0.6) | 116 (0.6) | 123 (0.7) | 124 (0.7) | 477 (0.6) | 0.876 |
| Time to bladder cancer, year | 17.0 (15.8, 17.9) | 17.0 (15.4, 17.9) | 17.0 (15.3, 18.0) | 16.9 (15.0, 18.0) | 17.0 (15.3, 18.0) | 0.056 |
| Liver cancer | 180 (1.0) | 155 (0.8) | 118 (0.6) | 114 (0.6) | 567 (0.8) | <0.001 |
| Time to civer cancer, year | 17.0 (15.9, 17.9) | 17.0 (15.4, 17.9) | 17.0 (15.3, 18.0) | 16.9 (15.0, 18.0) | 17.0 (15.3, 18.0) | 0.098 |

**Supplementary Table 3. The outcome of patients in the study cohorts of four quartiles time-weighted remnant cholesterol.**

Q: quantile; SD: standard deviation

| **Characteristics** | **Q1 (N=18894) Mean(SD);N or Count(%)** | **Q2 (N=18820) Mean(SD);N or Count(%)** | **Q3 (18757) Mean(SD);N or Count(%)** | **Q4 (N=18871) Mean(SD);N or Count(%)** | **Total (N=75342) Mean(SD);N or Count(%)** | **P value** |
|---|---|---|---|---|---|---|
| Overall cancer | 2191 (11.6) | 2047 (10.9) | 2048 (10.9) | 2049 (10.9) | 8335 (11.1) | 0.062 |
| Time to cancer, year | 16.8 (14.8, 17.9) | 17.0 (15.8, 17.9) | 17.0 (15.4, 18.0) | 16.2 (11.0, 17.8) | 16.8 (14.4, 17.9) | <0.001 |
| Lung cancer | 391 (2.1) | 424 (2.3) | 378 (2.0) | 447 (2.4) | 1640 (2.2) | 0.069 |
| Time to lung cancer, year | 17.0 (15.7, 17.9) | 17.1 (16.1, 18.0) | 17.1 (16.0, 18.0) | 16.3 (12.0, 17.9) | 17.0 (15.3, 18.0) | <0.001 |
| Gastrointestinal cancer | 1020 (5.4) | 965 (5.1) | 886 (4.7) | 919 (4.9) | 3790 (5.0) | 0.015 |
| Time to gastrointestinal cancer, year | 17.0 (15.3, 17.9) | 17.1 (16.0, 18.0) | 17.1 (16.0, 18.0) | 16.3 (11.7, 17.9) | 16.9 (15.2, 17.9) | <0.001 |
| Breast cancer | 320 (1.7) | 326 (1.7) | 391 (2.1) | 333 (1.8) | 1370 (1.8) | 0.017 |
| Time to Breast cancer, year | 17.0 (15.5, 17.9) | 17.1 (16.1, 18.0) | 17.1 (16.0, 18.0) | 16.3 (11.8, 17.9) | 16.9 (15.2, 17.9) | <0.001 |
| Ovarian cancer | 28 (0.1) | 24 (0.1) | 35 (0.2) | 36 (0.2) | 123 (0.2) | 0.356 |
| Time to ovarian cancer, year | 17.0 (15.8, 17.9) | 17.1 (16.1, 18.0) | 17.1 (16.0, 18.0) | 16.3 (12.1, 17.9) | 17.0 (15.3, 18.0) | <0.001 |
| Prostate cancer | 250 (1.3) | 217 (1.2) | 214 (1.1) | 181 (1.0) | 862 (1.1) | 0.011 |
| Time to prostate cancer, year | 17.0 (15.6, 17.9) | 17.1 (16.1, 18.0) | 17.1 (16.0, 18.0) | 16.3 (12.0, 17.9) | 16.9 (15.3, 18.0) | <0.001 |
| Genitourinary cancer | 562 (3.0) | 522 (2.8) | 534 (2.8) | 499 (2.6) | 2117 (2.8) | 0.266 |
| Time to genitourinary cancer, year | 17.0 (15.4, 17.9) | 17.1 (16.0, 18.0) | 17.1 (16.0, 18.0) | 16.3 (11.8, 17.9) | 16.9 (15.2, 17.9) | <0.001 |
| Colorectal cancer | 549 (2.9) | 508 (2.7) | 518 (2.8) | 476 (2.5) | 2051 (2.7) | 0.145 |
| Time to colorectal cancer, year | 17.0 (15.5, 17.9) | 17.1 (16.1, 18.0) | 17.1 (16.0, 18.0) | 16.3 (11.8, 17.9) | 16.9 (15.2, 17.9) | <0.001 |
| Pancreas cancer | 82 (0.4) | 70 (0.4) | 64 (0.3) | 76 (0.4) | 292 (0.4) | 0.507 |
| Time to pancreas cancer, year | 17.0 (15.8, 17.9) | 17.1 (16.1, 18.0) | 17.1 (16.0, 18.0) | 16.3 (12.1, 17.9) | 17.0 (15.3, 18.0) | <0.001 |
| Time to metastatic solid tumour, year | 124 (0.7) | 112 (0.6) | 118 (0.6) | 123 (0.7) | 477 (0.6) | 0.873 |
| Bladder cancer | 17.0 (15.7, 17.9) | 17.1 (16.1, 18.0) | 17.1 (16.0, 18.0) | 16.3 (12.0, 17.9) | 17.0 (15.3, 18.0) | <0.001 |
| Time to bladder cancer, year | 144 (0.8) | 150 (0.8) | 132 (0.7) | 141 (0.7) | 567 (0.8) | 0.77 |
| Liver cancer | 17.0 (15.8, 17.9) | 17.1 (16.1, 18.0) | 17.1 (16.0, 18.0) | 16.3 (12.1, 17.9) | 17.0 (15.3, 18.0) | <0.001 |

**Supplementary Figure 1 Distribution and association analysis between RC with time-weighted RC, HDL-C, LDL-C, TC, and TG.**

HDL-C = high density lipoprotein cholesterol, LDL-C = low density lipoprotein cholesterol, RR = risk ratios, TC = total cholesterol, TG = triglyceride, RC=remnant cholesterol.

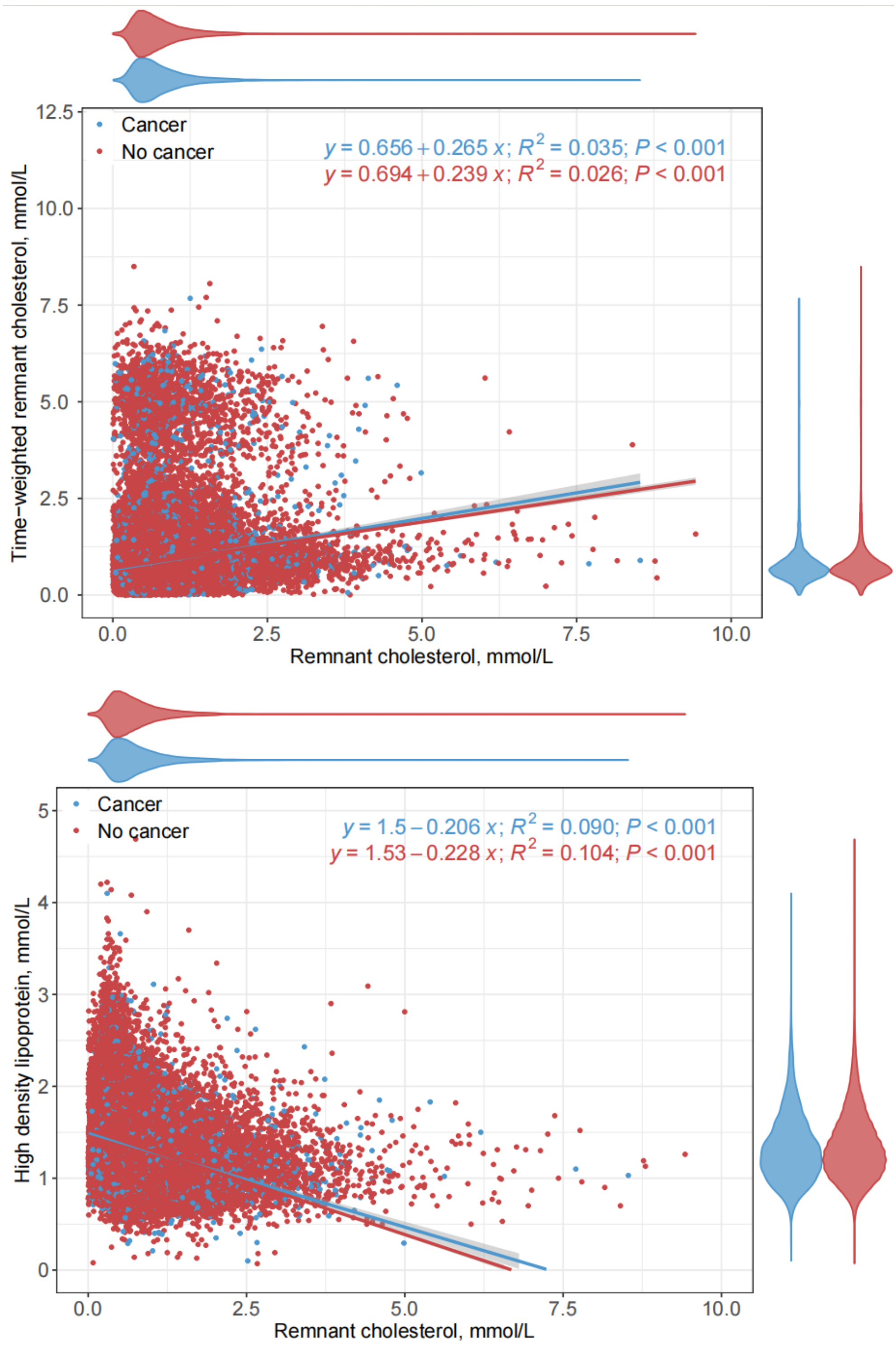

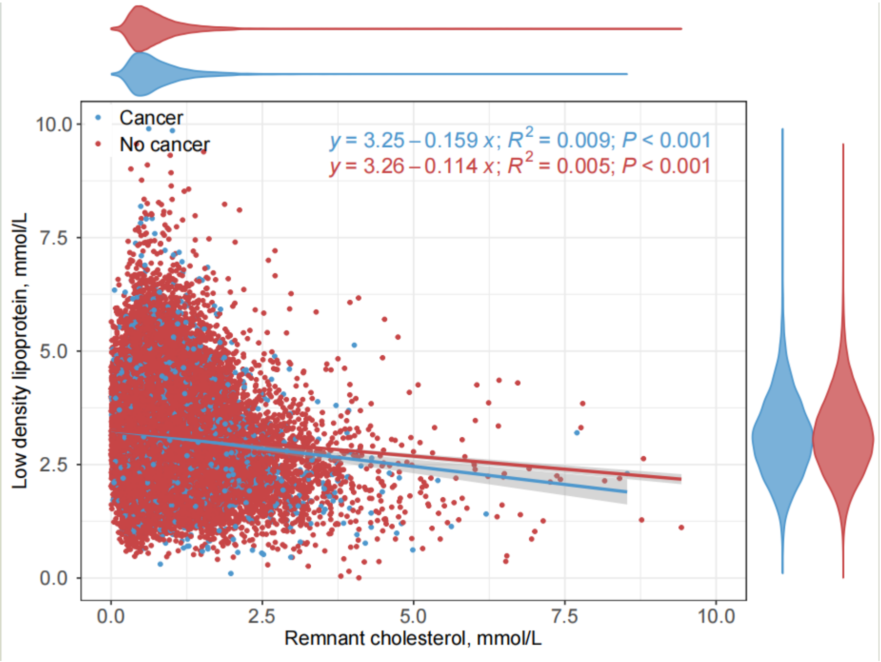
Cancer
No cancer
y = 3.25 – 0.159 x; R2 = 0.009; P < 0.001
y = 3.26 – 0.114 x; R2 = 0.005; P < 0.001
Low density lipoprotein, mmol/L
Remnant cholesterol, mmol/L
0.0
2.5
5.0
7.5
10.0

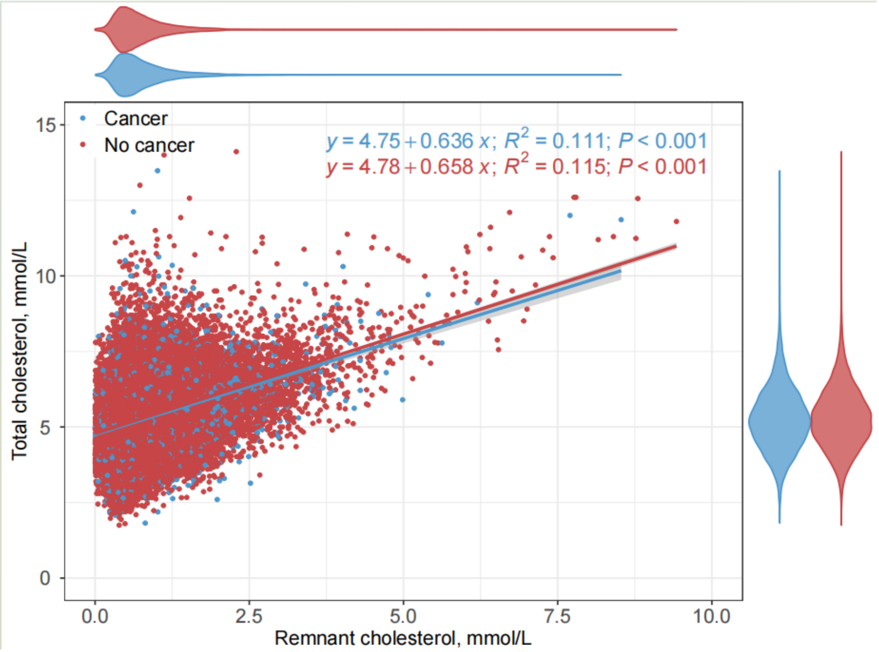
Cancer
No cancer
y = 4.75 + 0.636 x; R2 = 0.111; P < 0.001
y = 4.78 + 0.658 x; R2 = 0.115; P < 0.001
Total cholesterol, mmol/L
Remnant cholesterol, mmol/L
0
5
10
15
0.0
2.5
5.0
7.5
10.0

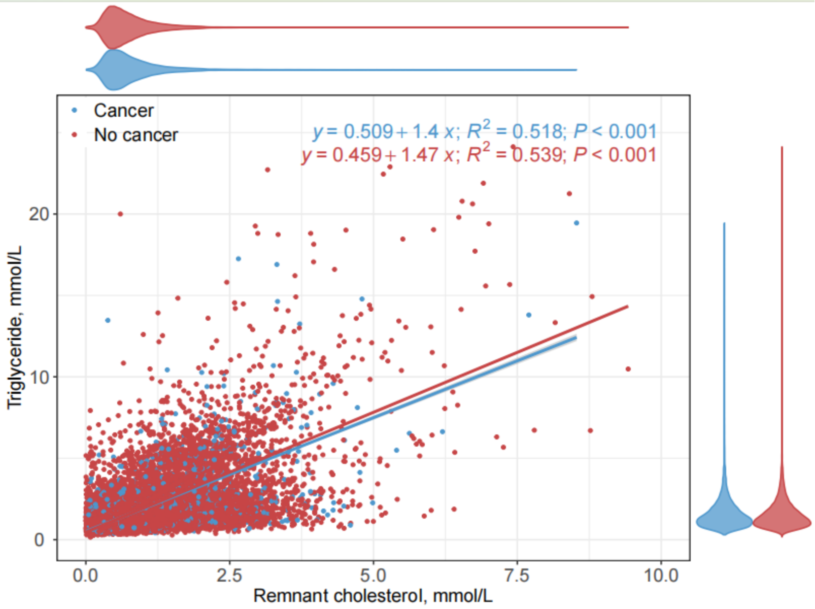

Cancer
No cancer
y = 0.509 + 1.4 x; R2 = 0.518; P < 0.001
y = 0.459 + 1.47 x; R2 = 0.539; P < 0.001
Triglyceride, mmol/L
Remnant cholesterol, mmol/L
0
10
20
0.0
2.5
5.0
7.5
10.0

**Supplementary Figure 2. Cumulative incidence curves stratified by quartiles of remnant cholesterol to predict overall and individual cancer.**

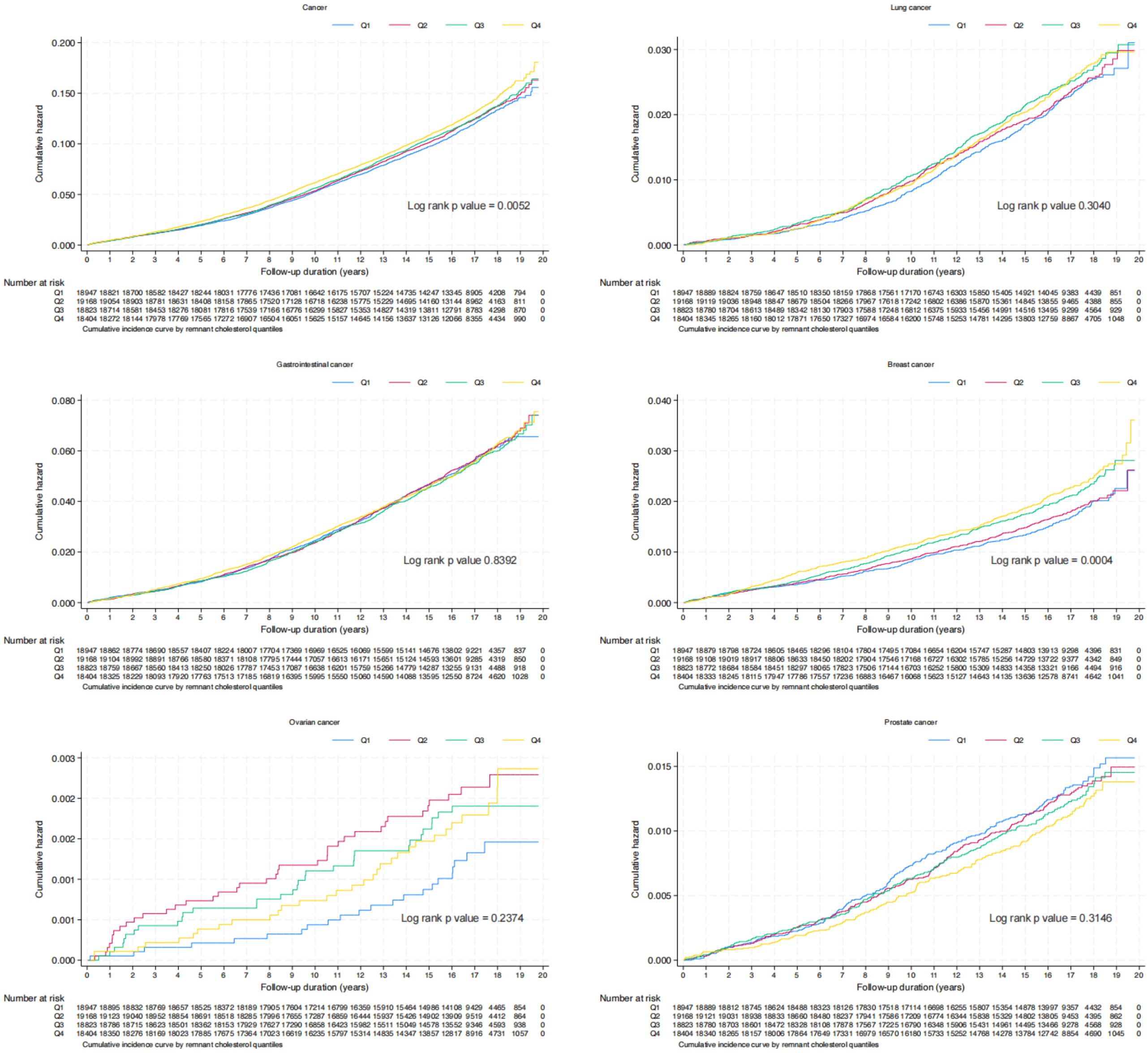

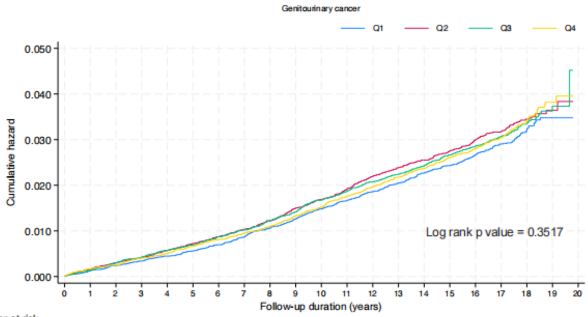


Number at risk

| | 0 | 1 | 2 | 3 | 4 | 5 | 6 | 7 | 8 | 9 | 10 | 11 | 12 | 13 | 14 | 15 | 16 | 17 | 18 | 19 | 20 |
|---|---|---|---|---|---|---|---|---|---|---|---|---|---|---|---|---|---|---|---|---|---|
| Q1 | 18947 | 18872 | 18788 | 18711 | 18581 | 18434 | 18256 | 18049 | 17745 | 17428 | 17021 | 16598 | 16150 | 15693 | 15232 | 14750 | 13865 | 9267 | 4386 | 844 | 0 |
| Q2 | 19168 | 19097 | 18993 | 18887 | 18766 | 18579 | 18386 | 18127 | 17820 | 17451 | 17064 | 16619 | 16179 | 15673 | 15169 | 14643 | 13645 | 9345 | 4353 | 855 | 0 |
| Q3 | 18823 | 18761 | 18667 | 18556 | 18409 | 18255 | 18017 | 17775 | 17457 | 17106 | 16659 | 16202 | 15757 | 15285 | 14811 | 14336 | 13311 | 9164 | 4513 | 913 | 0 |
| Q4 | 18404 | 18322 | 18234 | 18103 | 17943 | 17787 | 17561 | 17235 | 16878 | 16455 | 16055 | 15599 | 15103 | 14615 | 14122 | 13628 | 12587 | 8744 | 4628 | 1033 | 0 |

Cumulative incidence curve by remnant cholesterol quantiles

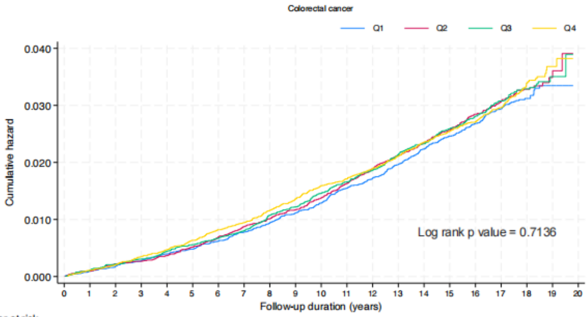


Number at risk

| | 0 | 1 | 2 | 3 | 4 | 5 | 6 | 7 | 8 | 9 | 10 | 11 | 12 | 13 | 14 | 15 | 16 | 17 | 18 | 19 | 20 |
|---|---|---|---|---|---|---|---|---|---|---|---|---|---|---|---|---|---|---|---|---|---|
| Q1 | 18947 | 18878 | 18798 | 18720 | 18591 | 18452 | 18279 | 18071 | 17774 | 17452 | 17053 | 16614 | 16169 | 15710 | 15246 | 14775 | 13895 | 9292 | 4394 | 840 | 0 |
| Q2 | 19168 | 19110 | 19011 | 18917 | 18806 | 18622 | 18423 | 18166 | 17863 | 17514 | 17130 | 16687 | 16252 | 15728 | 15211 | 14684 | 13692 | 9357 | 4351 | 855 | 0 |
| Q3 | 18823 | 18767 | 18681 | 18576 | 18434 | 18281 | 18058 | 17821 | 17498 | 17153 | 16702 | 16265 | 15819 | 15329 | 14855 | 14366 | 13337 | 9188 | 4514 | 928 | 0 |
| Q4 | 18404 | 18332 | 18241 | 18113 | 17950 | 17792 | 17555 | 17229 | 16866 | 16448 | 16052 | 15619 | 15131 | 14658 | 14161 | 13669 | 12632 | 8772 | 4647 | 1033 | 0 |

Cumulative incidence curve by remnant cholesterol quantiles

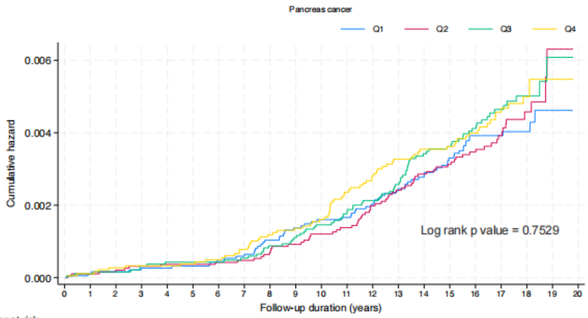


Number at risk

| | 0 | 1 | 2 | 3 | 4 | 5 | 6 | 7 | 8 | 9 | 10 | 11 | 12 | 13 | 14 | 15 | 16 | 17 | 18 | 19 | 20 |
|---|---|---|---|---|---|---|---|---|---|---|---|---|---|---|---|---|---|---|---|---|---|
| Q1 | 18947 | 18894 | 18829 | 18766 | 18654 | 18523 | 18366 | 18184 | 17899 | 17595 | 17206 | 16790 | 16348 | 15900 | 15457 | 14981 | 14103 | 9429 | 4466 | 856 | 0 |
| Q2 | 19168 | 19126 | 19046 | 18959 | 18863 | 18702 | 18530 | 18298 | 18009 | 17669 | 17300 | 16873 | 16455 | 15951 | 15439 | 14916 | 13923 | 9523 | 4416 | 864 | 0 |
| Q3 | 18823 | 18786 | 18719 | 18626 | 18504 | 18366 | 18156 | 17930 | 17628 | 17290 | 16863 | 16428 | 15988 | 15516 | 15048 | 14587 | 13560 | 9352 | 4596 | 939 | 0 |
| Q4 | 18404 | 18350 | 18274 | 18168 | 18022 | 17887 | 17677 | 17364 | 17019 | 16616 | 16233 | 15795 | 15313 | 14839 | 14352 | 13861 | 12821 | 8921 | 4735 | 1057 | 0 |

Cumulative incidence curve by remnant cholesterol quantiles

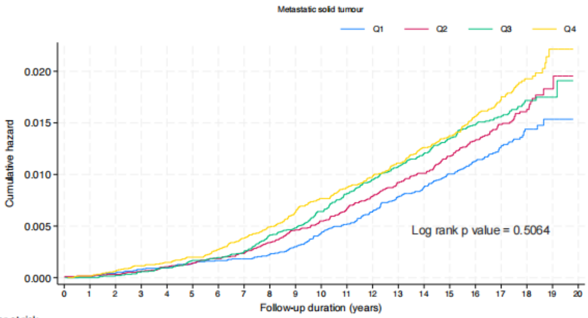


Number at risk

| | 0 | 1 | 2 | 3 | 4 | 5 | 6 | 7 | 8 | 9 | 10 | 11 | 12 | 13 | 14 | 15 | 16 | 17 | 18 | 19 | 20 |
|---|---|---|---|---|---|---|---|---|---|---|---|---|---|---|---|---|---|---|---|---|---|
| Q1 | 18947 | 18895 | 18829 | 18763 | 18651 | 18517 | 18364 | 18182 | 17895 | 17586 | 17194 | 16776 | 16334 | 15881 | 15441 | 14968 | 14086 | 9418 | 4459 | 855 | 0 |
| Q2 | 19168 | 19124 | 19046 | 18957 | 18855 | 18694 | 18522 | 18291 | 17992 | 17648 | 17279 | 16847 | 16429 | 15917 | 15408 | 14878 | 13885 | 9497 | 4407 | 863 | 0 |
| Q3 | 18823 | 18788 | 18719 | 18625 | 18502 | 18356 | 18150 | 17922 | 17614 | 17279 | 16848 | 16412 | 15974 | 15504 | 15041 | 14573 | 13543 | 9340 | 4590 | 939 | 0 |
| Q4 | 18404 | 18350 | 18272 | 18166 | 18017 | 17878 | 17662 | 17350 | 17001 | 16601 | 16216 | 15780 | 15297 | 14826 | 14333 | 13840 | 12797 | 8905 | 4730 | 1053 | 0 |

Cumulative incidence curve by remnant cholesterol quantiles

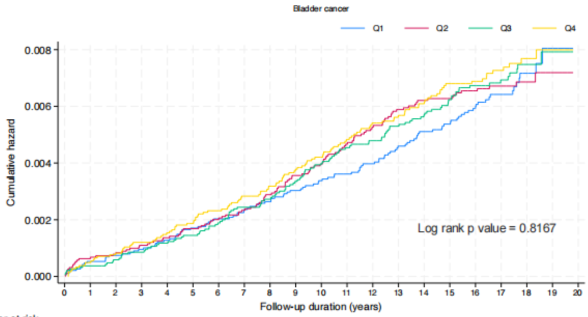


Number at risk

| | 0 | 1 | 2 | 3 | 4 | 5 | 6 | 7 | 8 | 9 | 10 | 11 | 12 | 13 | 14 | 15 | 16 | 17 | 18 | 19 | 20 |
|---|---|---|---|---|---|---|---|---|---|---|---|---|---|---|---|---|---|---|---|---|---|
| Q1 | 18947 | 18886 | 18818 | 18754 | 18637 | 18500 | 18340 | 18154 | 17870 | 17567 | 17179 | 16763 | 16319 | 15866 | 15420 | 14943 | 14066 | 9407 | 4451 | 852 | 0 |
| Q2 | 19168 | 19115 | 19033 | 18942 | 18841 | 18674 | 18497 | 18259 | 17965 | 17622 | 17254 | 16823 | 16407 | 15894 | 15388 | 14868 | 13877 | 9497 | 4412 | 862 | 0 |
| Q3 | 18823 | 18781 | 18710 | 18615 | 18489 | 18348 | 18130 | 17898 | 17596 | 17250 | 16813 | 16373 | 15940 | 15465 | 14999 | 14535 | 13508 | 9315 | 4578 | 936 | 0 |
| Q4 | 18404 | 18342 | 18262 | 18151 | 18000 | 17862 | 17646 | 17329 | 16983 | 16577 | 16190 | 15748 | 15266 | 14792 | 14305 | 13810 | 12775 | 8886 | 4720 | 1055 | 0 |

Cumulative incidence curve by remnant cholesterol quantiles

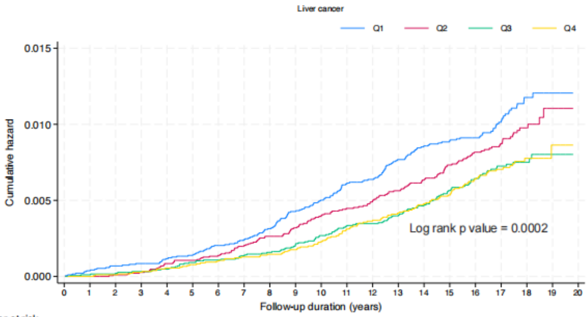


Number at risk

| | 0 | 1 | 2 | 3 | 4 | 5 | 6 | 7 | 8 | 9 | 10 | 11 | 12 | 13 | 14 | 15 | 16 | 17 | 18 | 19 | 20 |
|---|---|---|---|---|---|---|---|---|---|---|---|---|---|---|---|---|---|---|---|---|---|
| Q1 | 18947 | 18891 | 18823 | 18760 | 18645 | 18512 | 18354 | 18172 | 17888 | 17574 | 17181 | 16760 | 16323 | 15869 | 15423 | 14953 | 14084 | 9410 | 4449 | 853 | 0 |
| Q2 | 19168 | 19127 | 19048 | 18958 | 18853 | 18692 | 18516 | 18277 | 17984 | 17643 | 17274 | 16846 | 16435 | 15931 | 15417 | 14890 | 13896 | 9507 | 4412 | 863 | 0 |
| Q3 | 18823 | 18786 | 18717 | 18627 | 18504 | 18363 | 18151 | 17928 | 17626 | 17281 | 16846 | 16412 | 15979 | 15504 | 15035 | 14567 | 13540 | 9332 | 4588 | 935 | 0 |
| Q4 | 18404 | 18351 | 18276 | 18170 | 18021 | 17885 | 17672 | 17358 | 17017 | 16616 | 16230 | 15786 | 15304 | 14829 | 14335 | 13845 | 12802 | 8910 | 4730 | 1056 | 0 |

Cumulative incidence curve by remnant cholesterol quantiles

**Supplementary Figure 3. Cumulative incidence curves stratified by quartiles of time-weighted remnant cholesterol to predict overall and individual cancer.**

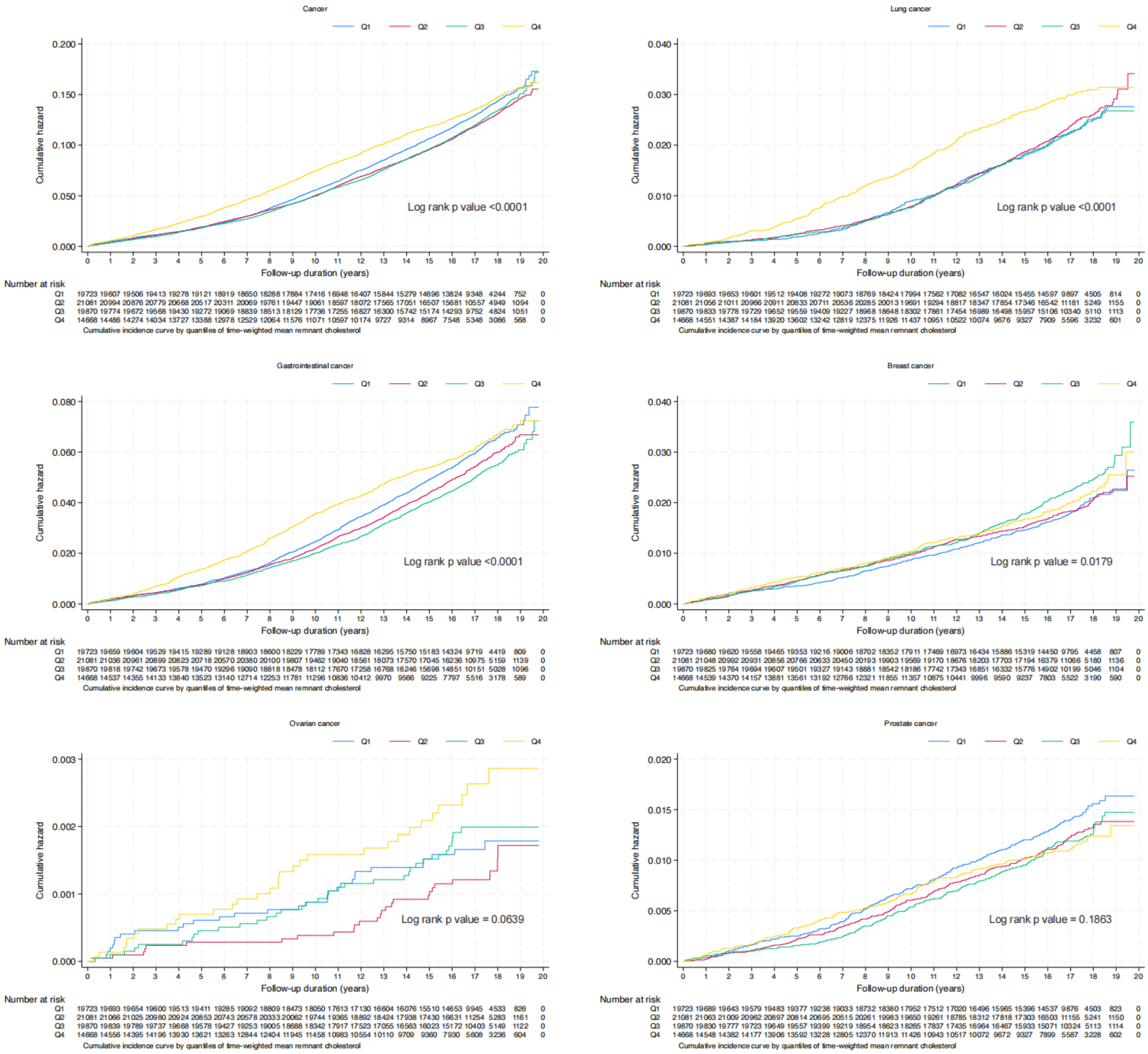

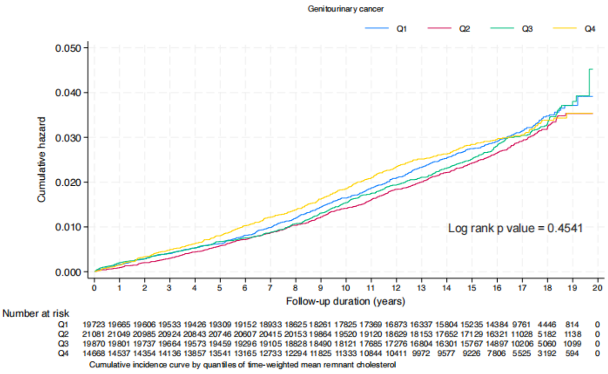
Genitourinary cancer
Q1 Q2 Q3 Q4
Cumulative hazard
0.050
0.040
0.030
0.020
0.010
0.000
Log rank p value = 0.4541
0 1 2 3 4 5 6 7 8 9 10 11 12 13 14 15 16 17 18 19 20
Follow-up duration (years)
Number at risk
Q1 19723 19665 19606 19533 19426 19309 19152 18933 18625 18261 17825 17369 16873 16337 15804 15235 14384 9761 4446 814 0
Q2 21081 21049 20985 20924 20843 20746 20607 20415 20153 19864 19520 19120 18629 18153 17652 17129 16321 11028 5182 1138 0
Q3 19870 19801 19737 19664 19573 19459 19296 19105 18828 18490 18121 17685 17276 16804 16301 15767 14897 10206 5060 1099 0
Q4 14668 14537 14354 14136 13857 13541 13165 12733 12294 11825 11333 10844 10411 9972 9577 9226 7806 5525 3192 594 0
Cumulative incidence curve by quantiles of time-weighted mean remnant cholesterol

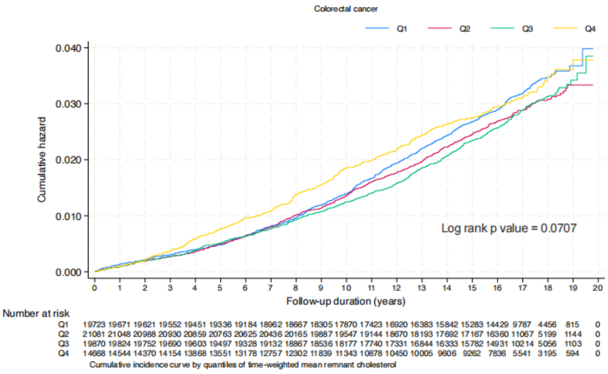
Colorectal cancer
Q1 Q2 Q3 Q4
Cumulative hazard
0.040
0.030
0.020
0.010
0.000
Log rank p value = 0.0707
0 1 2 3 4 5 6 7 8 9 10 11 12 13 14 15 16 17 18 19 20
Follow-up duration (years)
Number at risk
Q1 19723 19671 19621 19552 19451 19336 19184 18962 18667 18305 17870 17423 16920 16383 15842 15283 14429 9787 4456 815 0
Q2 21081 21048 20988 20930 20859 20763 20625 20436 20165 19887 19547 19144 18670 18193 17692 17167 16360 11067 5199 1144 0
Q3 19870 19824 19752 19690 19603 19497 19328 19132 18867 18536 18177 17740 17331 16844 16333 15782 14931 10214 5056 1103 0
Q4 14668 14544 14370 14154 13868 13551 13178 12757 12302 11839 11343 10878 10450 10005 9606 9262 7836 5541 3195 594 0
Cumulative incidence curve by quantiles of time-weighted mean remnant cholesterol

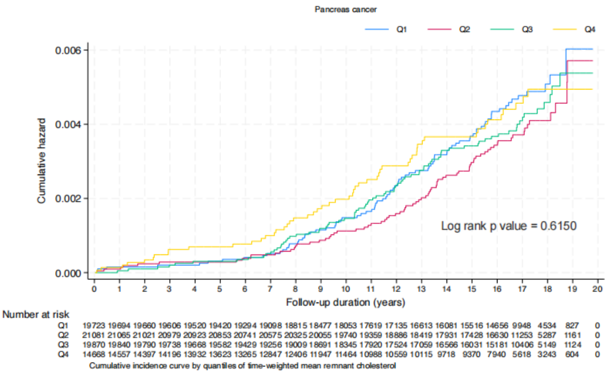
Pancreas cancer
Q1 Q2 Q3 Q4
Cumulative hazard
0.006
0.004
0.002
0.000
Log rank p value = 0.6150
0 1 2 3 4 5 6 7 8 9 10 11 12 13 14 15 16 17 18 19 20
Follow-up duration (years)
Number at risk
Q1 19723 19694 19660 19606 19520 19420 19294 19098 18815 18477 18053 17619 17135 16613 16081 15516 14656 9948 4534 827 0
Q2 21081 21065 21021 20979 20923 20853 20741 20575 20325 20055 19740 19359 18886 18419 17931 17428 16630 11253 5287 1161 0
Q3 19870 19840 19790 19738 19668 19582 19429 19256 19009 18691 18345 17920 17524 17059 16566 16031 15181 10406 5149 1124 0
Q4 14668 14557 14397 14196 13932 13623 13265 12847 12406 11947 11464 10988 10559 10115 9718 9370 7940 5618 3243 604 0
Cumulative incidence curve by quantiles of time-weighted mean remnant cholesterol

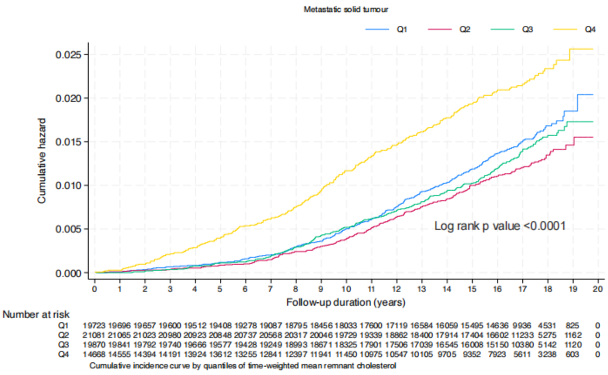
Metastatic solid tumour
Q1 Q2 Q3 Q4
Cumulative hazard
0.025
0.020
0.015
0.010
0.005
0.000
Log rank p value <0.0001
0 1 2 3 4 5 6 7 8 9 10 11 12 13 14 15 16 17 18 19 20
Follow-up duration (years)
Number at risk
Q1 19723 19696 19657 19600 19512 19408 19278 19087 18795 18456 18033 17600 17119 16584 16059 15495 14636 9936 4531 825 0
Q2 21081 21065 21023 20980 20923 20848 20737 20568 20317 20046 19729 19339 18862 18400 17914 17404 16602 11233 5275 1162 0
Q3 19870 19841 19792 19740 19666 19577 19428 19249 18993 18671 18325 17901 17506 17039 16545 16008 15150 10380 5142 1120 0
Q4 14668 14555 14394 14191 13924 13612 13255 12841 12397 11941 11450 10975 10547 10105 9705 9352 7923 5611 3238 603 0
Cumulative incidence curve by quantiles of time-weighted mean remnant cholesterol

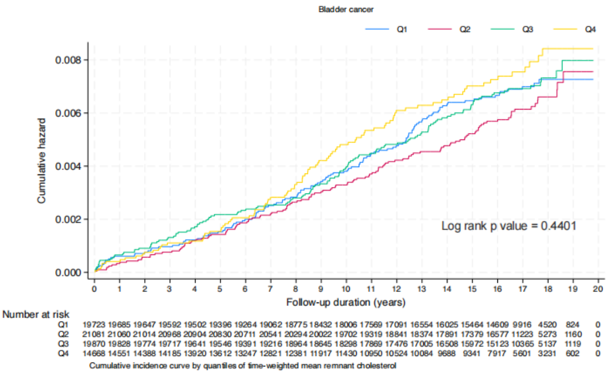
Bladder cancer
Q1 Q2 Q3 Q4
Cumulative hazard
0.008
0.006
0.004
0.002
0.000
Log rank p value = 0.4401
0 1 2 3 4 5 6 7 8 9 10 11 12 13 14 15 16 17 18 19 20
Follow-up duration (years)
Number at risk
Q1 19723 19685 19647 19592 19502 19396 19264 19062 18775 18432 18006 17569 17091 16554 16025 15464 14609 9916 4520 824 0
Q2 21081 21060 21014 20968 20904 20830 20711 20541 20294 20022 19702 19319 18841 18374 17891 17379 16577 11223 5273 1160 0
Q3 19870 19828 19774 19717 19641 19546 19391 19216 18964 18645 18298 17869 17476 17005 16508 15972 15123 10365 5137 1119 0
Q4 14668 14551 14388 14185 13920 13612 13247 12821 12381 11917 11430 10950 10524 10084 9688 9341 7917 5601 3231 602 0
Cumulative incidence curve by quantiles of time-weighted mean remnant cholesterol

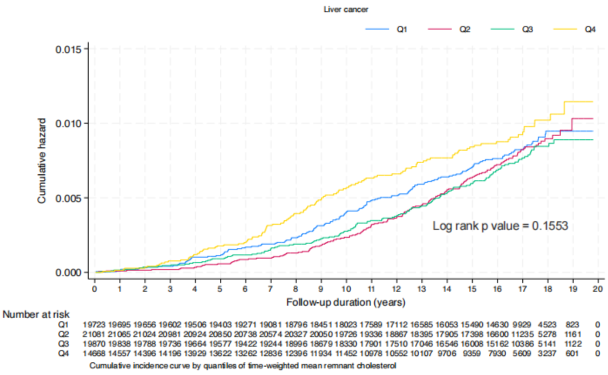
Liver cancer
Q1 Q2 Q3 Q4
Cumulative hazard
0.015
0.010
0.005
0.000
Log rank p value = 0.1553
0 1 2 3 4 5 6 7 8 9 10 11 12 13 14 15 16 17 18 19 20
Follow-up duration (years)
Number at risk
Q1 19723 19695 19656 19602 19506 19403 19271 19081 18796 18451 18023 17589 17112 16585 16053 15490 14630 9929 4523 823 0
Q2 21081 21065 21024 20981 20924 20850 20738 20574 20327 20050 19726 19336 18867 18395 17905 17398 16600 11235 5278 1161 0
Q3 19870 19838 19788 19736 19664 19577 19422 19244 18996 18679 18330 17901 17510 17046 16546 16008 15162 10386 5141 1122 0
Q4 14668 14557 14396 14196 13929 13622 13262 12836 12396 11934 11452 10978 10552 10107 9706 9359 7930 5609 3237 601 0
Cumulative incidence curve by quantiles of time-weighted mean remnant cholesterol

**Supplementary Figure 4. Forest plot of subgroup analyses between baseline RC with cancer**

RC: remnant cholesterol, HR: hazard ratio, CI: confidence interval.

| Variable | Count (%) | Hazard ratio (95% CI) | P for interaction |
|---|---|---|---|
| **Gender** | | | |
| Male | 29905 (39.7%) | 0.96 [0.90−1.02] | <0.001 |
| Female | 45437 (60.3%) | 1.17 [1.11−1.22] | |
| **Age** | | | |
| >65 | 32825 (43.6%) | 1.06 [1.00−1.11] | 0.371 |
| <65 | 42517 (56.4%) | 1.09 [1.04−1.15] | |
| **Cormorbidity** | | | |
| Dyslipidemia (Yes) | 1288 (1.7%) | 0.80 [0.63−1.01] | 0.011 |
| Dyslipidemia (No) | 74054 (98.3%) | 1.08 [1.04−1.12] | |
| Diabetes mellitus (Yes) | 7889 (10.5%) | 0.98 [0.89−1.08] | 0.086 |
| Diabetes mellitus (No) | 67453 (89.5%) | 1.08 [1.03−1.12] | |
| Hypertension (Yes) | 39026 (51.8%) | 1.04 [0.99−1.10] | 0.168 |
| Hypertension (No) | 36316 (48.2%) | 1.09 [1.04−1.15] | |
| Stroke/TIA (Yes) | 508 (0.7%) | 0.71 [0.43−1.17] | 0.096 |
| Stroke/TIA (No) | 74834 (99.3%) | 1.08 [1.04−1.12] | |
| IHD (Yes) | 1516 (2.0%) | 0.99 [0.78−1.26] | 0.471 |
| IHD (No) | 73826 (98.0%) | 1.08 [1.04−1.12] | |
| Liver diseases (Yes) | 269 (0.4%) | 0.95 [0.49−1.84] | 0.732 |
| Liver diseases (No) | 75073 (99.6%) | 1.08 [1.04−1.12] | |
| Renal diseases (Yes) | 105 (0.1%) | 2.08 [0.67−6.49] | 0.298 |
| Renal diseases (No) | 75237 (99.9%) | 1.08 [1.04−1.11] | |
| **Medication use** | | | |
| ACEI/ARB (Yes) | 8890 (11.8%) | 1.04 [0.94−1.15] | 0.534 |
| ACEI/ARB (No) | 66452 (88.2%) | 1.07 [1.03−1.12] | |
| Antiplatelets (Yes) | 7921 (10.5%) | 1.00 [0.90−1.11] | 0.156 |
| Antiplatelets (No) | 67421 (89.5%) | 1.08 [1.04−1.12] | |
| Lipid lowering drugs (Yes) | 7021 (9.3%) | 0.98 [0.89−1.08] | 0.067 |
| Lipid lowering drugs (No) | 68321 (90.7%) | 1.08 [1.04−1.13] | |
| Statins and fibrates (Yes) | 8460 (11.2%) | 0.96 [0.88−1.05] | 0.004 |
| Statins and fibrates (No) | 66882 (88.8%) | 1.10 [1.06−1.15] | |
| Beta blockers (Yes) | 15006 (19.9%) | 1.09 [1.02−1.18] | 0.542 |
| Beta blockers (No) | 60336 (80.1%) | 1.07 [1.02−1.11] | |
| Non−steroidal anti−inflammatory drugs (Yes) | 7594 (10.1%) | 1.02 [0.91−1.13] | 0.29 |
| Non−steroidal anti−inflammatory drugs (No) | 67748 (89.9%) | 1.08 [1.04−1.12] | |
| Antidiabetic drugs (Yes) | 15121 (20.1%) | 1.05 [0.97−1.14] | 0.61 |
| Antidiabetic drugs (No) | 60221 (79.9%) | 1.08 [1.03−1.12] | |

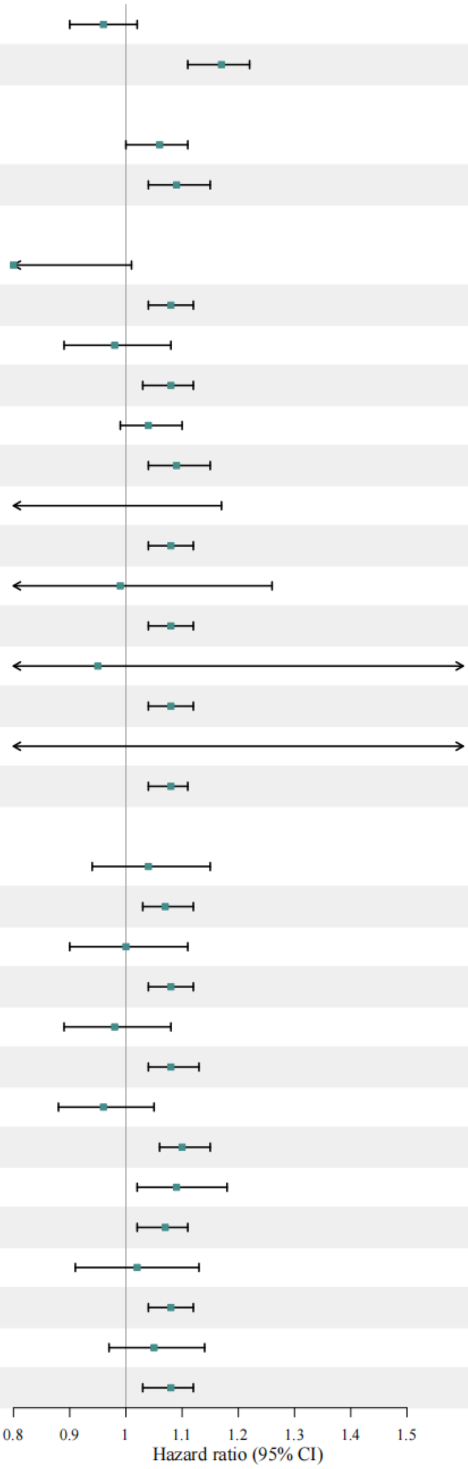
